# Using clustering of genetic variants in Mendelian randomization to interrogate the causal pathways underlying multimorbidity

**DOI:** 10.1101/2023.03.18.23287164

**Authors:** Xiaoran Liang, Ninon Mounier, Nicolas Apfel, Sara Khalid, Timothy M Frayling, Jack Bowden, GEMINI Consortium

## Abstract

Mendelian randomization (MR) is an epidemiological approach that utilizes genetic variants as instrumental variables to estimate the causal effect of a modifiable but likely confounded exposure on a health outcome. This paper investigates an MR scenario in which different subsets of genetic variants identify different causal effects. These variants may aggregate into clusters, and such variant clusters are likely to emerge if they affect the exposure and outcome via distinct biological pathways. In the framework of multi-outcome MR, where a common risk factor causally impacts several disease outcomes simultaneously, these variant clusters can reflect the heterogeneous effects this shared risk factor concurrently exerts on all the diseases under examination. This, in turn, can provide insights into the disease-causing mechanisms underpinning the co-occurrence of multiple long-term conditions, a phenomenon known as multimorbidity. To identify such variant clusters, we adapt the general method of Agglomerative Hierarchical Clustering (AHC) to the summary data MR setting, enabling cluster detection based on the variant-specific causal estimates, using only genome-wide summary statistics. In particular, we tailor the method for multi-outcome MR to aid the elucidation of the potentially multifaceted causal pathways underlying multimorbidity stemming from a shared risk factor. We show in various Monte Carlo simulations that our ‘MR-AHC’ method detects variant clusters with high accuracy, outperforming the existing multi-dimensional clustering methods. In an application example, we use the method to analyze the causal effects of high body fat percentage on a pair of well-known multimorbid conditions, type 2 diabetes (T2D) and osteoarthritis (OA), discovering distinct variant clusters reflecting heterogeneous causal effects. Pathway analyses of these variant clusters indicate interconnected cellular processes underlying the co-occurrence of T2D and OA; while the protective effect of higher adiposity on T2D could possibly be linked to the enhanced activity of ion channels related to insulin secretion.

## Introduction

Mendelian randomization (MR) is a widely used method in epidemiology that leverages genetic variants (usually in the form of single nucleotide polymorphisms, SNPs) as instrumental variables (IV) for estimating the causal effect of a potentially confounded exposure on an outcome [1, 2]. If a genetic variant is sufficiently associated with the exposure, independent of possible confounders of the exposure-outcome relationship, and affects the outcome only through the exposure, then it is a valid instrument for assessing causality [3]. With further parametric assumptions, for example that relationships between all variables are additive and linear, and all variants included as instruments encode a single homogeneous causal effect from the exposure to the outcome, then the causal parameter of interest can be estimated using simple meta-analytic methods based on genome-wide summary statistics [4–6]. In this setting, all the variant-specific causal estimates are expected to target the same, true causal effect, and their ‘ratio’ estimates (derived as the ratio of the variant-outcome to variant-exposure association), from which the overall meta-analysis is performed, should vary by sampling error alone [7, 8]. Excess ‘heterogeneity’ amongst the ratio estimates is therefore a sign that one or more of the assumptions has been violated [9].

A major source of excess heterogeneity is undoubtedly *horizontal pleiotropy*, the phenomenon whereby a variant affects multiple traits and therefore is associated with the outcome through pathways other than via the exposure [9, 10]. This has been extensively studied with improved methods for pleiotropy detection [8, 11] and robust estimation [5, 12, 13]. Violation of the causal effect homogeneity assumption, has, by contrast, been far less researched, despite this being a plausible feature of many analyses. For example, it is suspected that general adiposity, which is often proxied by a single trait like body mass index (BMI), exerts a heterogeneous causal effect on type 2 diabetes (T2D) depending on the location of the adipose tissue in the body (e.g. if it is peripheral or visceral) [14]. In this case, variants associated with different physiological aspects of the exposure may target distinct causal effects.

In the presence of excess heterogeneity from both sources, the genetic variants can be grouped into distinct clusters, such that all variants in each cluster indicate the same effect. Several studies have explored variant clusters in the MR framework. It is well recognized that it is impossible to discern whether each cluster embodies genuine causal mechanisms between the exposure and outcome, or is formed due to pleiotropic pathways, without further domain knowledge or modelling assumptions. Therefore, overdispersion caused by both sources can be summarized under an umbrella term such as “clustered heterogeneity”, as proposed by Foley et al. [15], or “mechanistic heterogeneity” by Iong et al. [16].

In this paper, we propose a method to identify variant clusters under mechanistic heterogeneity, building upon the Agglomerative Hierarchical Clustering (AHC) method developed by Apfel and Liang [17] in the field of econometrics for IV selection. We adapt the method to the summary-data MR setting, hence referring to it as “MR-AHC”, to group variants based on their ratio estimates using genome-wide summary statistics. More notably, we have tailored the method to the multi-outcome MR setting, in which a shared exposure causally impacts several disease outcomes simultaneously. This extension is specially crafted for investigating the causal mechanisms underpinning multimorbidity, which refers to the co-existence of two or more long-term conditions in one individual [18].

A substantial and growing proportion of the adult population is affected by multimorbidity, and it has been recognized as a global priority for health research [19, 20]. It is therefore important to comprehend the underlying disease-causing pathways. Numerous studies have identified common risk factors associated with a broad range of conditions. New methods have also been introduced for identifying these factors [21]. However, such shared risk factors are often complex traits and may exert heterogeneous influences on diseases through multifaceted mechanisms. For example, obesity, one of the most well-established risk factors contributing to various forms of multimorbidity [22, 23], is recognized to impact diseases through a variety of distinct pathways [14, 24].

Given the multitude of potential causal pathways stemming from a common risk factor, particularly in the case of complex traits like obesity, to enable effective clinical prevention and intervention, it is necessary to elucidate the mechanisms through which this common risk factor induces the co-occurrence of the conditions. A starting point can be identifying the variant clusters associated with diverse causal effects within a multi-outcome MR framework. To illustrate this, consider the hypothetical example depicted in Figure 1. Here, variants linked to the exposure *X* are divided into three groups (*G*_1_ to *G*_3_), as they influence the two disease outcomes *Y*_1_ and *Y*_2_ through three different aspects of the exposure *X* (denoted by *X*_1_ to *X*_3_) that might not be easy to measure directly. Among the three groups, *G*_1_ is associated with an increasing effect on *Y*_1_ but a protective effect against *Y*_2_, and *G*_3_ indicates an increasing effect on *Y*_2_ but no effect on *Y*_1_. Only the group *G*_2_ corresponds to pathways through which the shared risk factor increases the risk of both diseases. Therefore, identifying variant clusters can unveil the potentially heterogeneous causal effects, and subsequently shed light on the mechanisms linking the common risk factor to the co-occurrence of the conditions.

**Figure 1.**
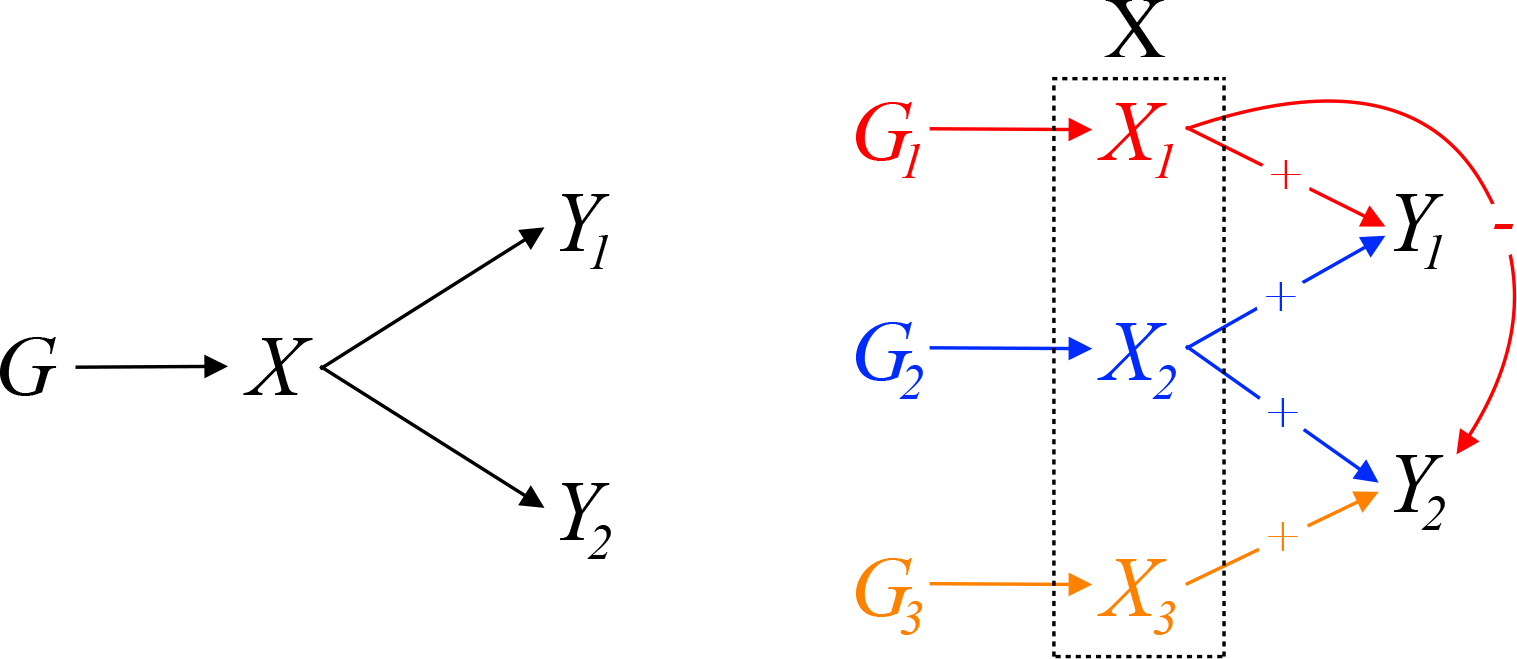
Left: Multi-outcome MR involving two disease outcomes and a common risk factor; right: clusters formed by the variants associated with the common exposure, which reflect heterogeneous causal pathways.

Several clustering approaches have been proposed within the MR framework to group genetic variants based on their causal estimates, such as MR-Clust [15] and MR-PATH [16]. However, these methods are primarily tailored to settings involving a single exposure and a single outcome, making them less suitable for handling the complexities of multimorbidity, as they lack multi-dimensional clustering options. On the other hand, methods such as NAvMix [25] do allow for multi-dimensional clustering of genetic variants, but are not inherently rooted in the MR framework, since they group variants based on their direct variant-trait associations, rather than causal estimates. This may limit their utility for causal inference. The mclust method [26] does permit multi-dimensional clustering using causal estimates, but we show that the method’s accuracy can be sub-optimal. In contrast, our MR-AHC method allows for multi-dimensional causal clustering based on MR estimates, whilst achieving a high clustering accuracy, which we have demonstrated in extensive Monte Carlo simulations.

We apply MR-AHC to investigate the causal effects of body fat percentage (BFP), as a shared risk factor, on a pair of multimorbid conditions, T2D and osteoarthritis (OA). Our analysis identifies four variant clusters indicating heterogeneous effects on both conditions. To provide insights into the underlying causal pathways, we conducted comprehensive gene-set analyses on the clusters, combining evidence from both canonical pathway analyses and gene-set Phenome-wide association analyses (PheWAS). While the clustering results cannot directly label a cluster as signifying genuine mechanisms or pleiotropic pathways, we show how post-clustering analyses may enable this distinction. Our findings on the cluster associated with increasing risks of both conditions indicate shared pathways underpinning the co-occurrence of T2D and OA through interconnected cellular processes related to gene expression transcription and cellular responses to stimuli. We provide further evidence using cluster-specific MR for the unifying pathway from obesity to the T2D-OA multimorbidity through elevated oxidative stress. Another cluster exhibits a protective effect against T2D, with integrated canonical pathway and PheWAS evidence supporting a possible mechanism involving enhanced activity of the ion channels related to insulin secretion. This might be linked to elevated levels of high-density lipoprotein cholesterol (HDL-C) associated with smaller waist-to-hip ratios.

## Results

### The MR-AHC method for clustering genetic variants in summary-data Mendelian randomization

We assume the following summary statistics for *J* genetic variants involved in an MR investigation: the variant-exposure association estimate 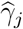, and the variant-outcome association estimate 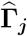, where *j* = 1, …, *J*. In an MR setting with a common exposure and multiple *P* outcomes, 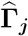 is the vector of the *P* variant-outcome associations. We maintain the assumption that the variant-exposure associations are measured from a sample independent from all the variant-outcome association samples, but overlap between the outcome samples is allowed. We also assume that all the genetic variants are themselves mutually uncorrelated (i.e. not in linkage disequilibrium). The causal estimates of the exposure on the *P* outcomes using only variant *j* as instrument (i.e. the ratio estimates) can then be obtained as the *P* -dimensional vector 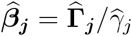. Let 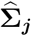 be an estimate of the covariance matrix of 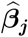.

We propose MR-AHC, a two-step procedure with 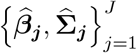 as inputs, to group genetic variants indicating the same causal effects, or in other words, having similar observed ratio estimates 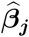, into the same cluster. We illustrated the method with a simple hypothetical example, shown in Figure 2. For ease of illustration, we consider the case with a single outcome, but the same procedure applies generally with multiple outcomes. The first step of the method, the merging step, is illustrated in the left panel of Figure 2. It shows a situation with six variants that form three clusters (one of them comprised of a single variant). The dotted lines at *β*_1_ and *β*_2_ are the true heterogeneous causal effects from the exposure to the outcome, and the circles above the real line denote the variant-specific ratio estimates. The differences in the size of the circles reflect the fact that summary data estimates exhibit varying degrees of uncertainty. In the explanation below, we refer to these estimates and their corresponding variants by the numbers 1 to 6, from left to right.

**Figure 2.**
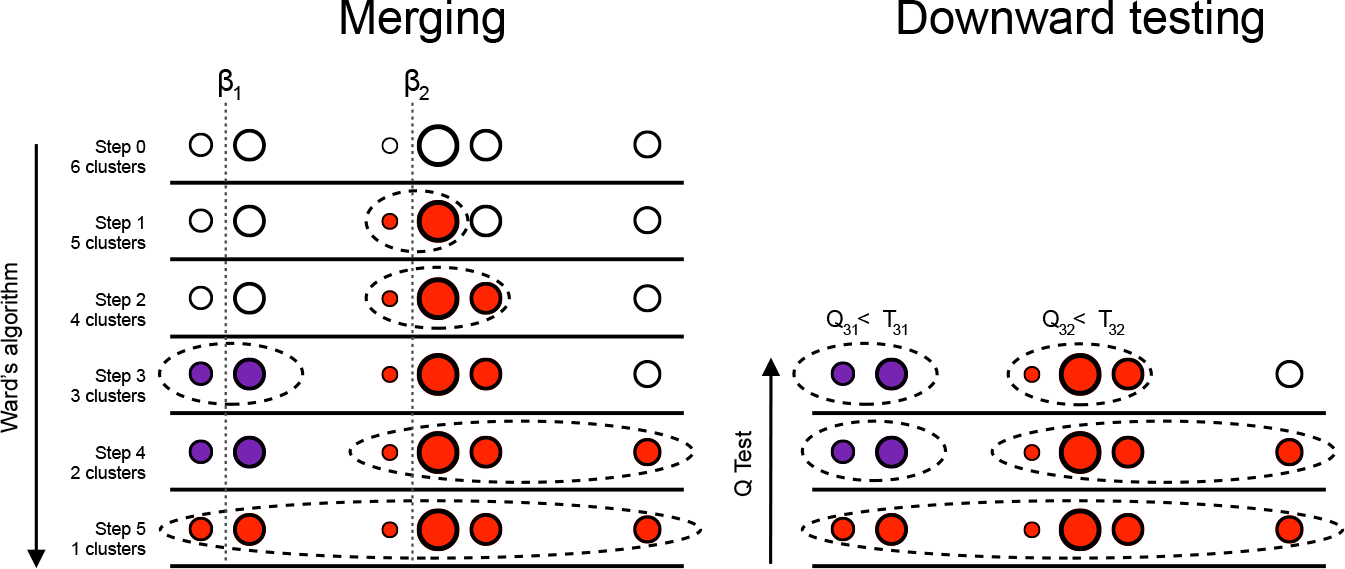
Illustration of the MR-AHC method for a hypothetical example adapted from Apfel and Liang [17]. Left panel: Ward’s algorithm defines a clustering path. Right panel: A downward testing procedure is applied until the step-specific heterogeneity statistics can not be rejected at a specified threshold.

In the initialization step of the merging process (Step 0 in the illustration), each variant-specific estimate has its own cluster. Next, we merge the two estimates which are closest in terms of their weighted squared Euclidean distance, i.e. those estimated with Variant 3 and 4 (the two red circles). These two estimates are merged into one cluster and we now have five clusters left. We re-calculate the pairwise distances with the five clusters and merge the closest two into a new cluster. We continue with this procedure until step 5 where all variants are in a single cluster.

By the end of the merging step, we have generated a clustering path. Along each step of the path, the number of clusters, denoted by *K*, varies from *K* = 1 to *K* = *J* by increments of 1. Next, in the second step of MR-AHC, we re-trace the clustering path to select the optimal value of *K* using a downward testing procedure, operates as follows: starting from the largest cluster containing all variants, apply Cochran’s Q test [27] to examine the degree of heterogeneity of all the ratio estimates by calculating the test statistic and comparing it with a pre-specified significance threshold. If the null hypothesis of “no excess heterogeneity” gets rejected, then move to the next level of the clustering path and apply the Q test to all the sub-clusters on that level. We repeat this process until reaching a level where no sub-cluster heterogeneity statistic rejects at the given significance threshold. In our illustrative example, we would expect the downward testing procedure to re-trace from step 5 to step 3 of the clustering path, thus determining three groups formed by Variants 1-2, Variants 3-5, and Variant 6 alone.

In the original AHC algorithm proposed by Apfel and Liang [17], the inputs are essentially just the ratio estimates 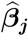, hence the clustering objects are treated as non-random fixed data points. MR-AHC adjusts the algorithm to take into account the uncertainty of 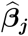 by incorporating the covariance matrix 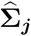 into the weighted squared Euclidean distance in the merging process. We show in Appendix A that this distance between two clusters is essentially the Wald statistic for testing the null hypothesis that “the two clusters indicate the same causal effect”. Therefore, merging two clusters with the smallest distance can be interpreted as merging two clusters with the highest similarity in their cluster-specific causal effects.

In terms of the covariance matrix estimate 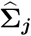, we show in Appendix A that if all the outcome samples are non-overlapping and/or the phenotypic correlations between the outcome traits are zero, then all the ratio estimates of a given variant are uncorrelated. In this case, all the covariance terms are zero and 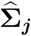 is just a diagonal matrix with the non-zero entries being the variances of the ratio estimates, which can be easily estimated from the GWAS summary statistics. If the covariances are non-zero, we show that 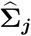 can be estimated via linkage disequilibrium (LD) score regression [28] and seamlessly incorporated into the analysis.

In the downward testing procedure, following the recommendation in Belloni et al. [29], we define the threshold p-value for the Q test as ζ= 0.1*/* log(*n*) where *n* is the sample size. We prove in Appendix B that this threshold p-value results in a consistent clustering procedure. That is, as *n* increases, the probability of correctly identifying all true members of each cluster tends to 1. If the exposure and outcome samples are of different sizes, we recommend using the sample size of the smallest outcome sample. For binary outcomes, an effective sample size can be approximated with the number of cases and controls, see Han and Eskin [30].

MR-AHC does not require pre-specification of the number of clusters. It can also easily identify a “null cluster” and a “junk cluster”, following the terminology of Foley et al. [15], which refer to, respectively, the cluster identifying a zero causal effect, and the cluster containing variants not assigned to any detected clusters. Specifically, we conduct a post-clustering Wald test on each cluster-specific causal estimate for the null hypothesis of a zero causal effect using ζ= 0.1*/* log(*n*) as the threshold significance p-value. For the junk cluster, we simply classify all variants that do not fit into any other clusters as junk variants. To further improve the clustering accuracy of MR-AHC, we also extend the basic algorithm illustrated above to an outlier-robust version, to correct for the outliers in the ratio estimates, see the method section for details.

### Simulation results

We conduct Monte Carlo simulations to evaluate the performance of the MR-AHC method in detecting variant clusters and estimating the causal effects in various settings that mimic the multimorbidity scenarios we are interested in, which involve a shared exposure causally affecting multiple outcome conditions. We consider 12 simulation designs, where the number of outcomes is either *P* = 2 or *P* = 3, the number of substantive variant clusters is either *K* = 1 or *K* = 4, and the sample correlation between the outcomes is either *ρ* = 0, *ρ* = 0.2 or *ρ* = 0.7 (see the method section for a detailed definition of *ρ*). In all designs, we have *J* = 100 SNPs with 10 designated as true ‘junk’ variants.

The two classes of scenarios stratified by the number of variant clusters are illustrated in Figure 3. The directed acyclic graph (DAG) in Panel (a) illustrates the data generation process when there are four substantive clusters and one noise cluster. Multiple outcomes (two or three) are represented by the single notation *Y*. Variant clusters are formed due to differential sub-components of the exposure, denoted by *X*_1_ to *X*_5_. Variant Cluster 1 and *X*_1_ represent a correlated pleiotropy pathway, and Clusters 2 to 4 correspond to genuine heterogeneous causal mechanisms from the exposure to the outcomes. The scatter plot on the right of Panel (a) is based on a representative simulated dataset of the two-outcome case. We also examine the performance of the method when there is actually no mechanistic heterogeneity, i.e. there is only one real cluster and one noise cluster. The design is shown by a DAG and representative dataset in Figure 3 Panel (b). The arrow from *X*_1_ to *Y* is absent, meaning that the only substantive cluster is also a null cluster and there is no causal effect between the exposure and the outcomes. See the method section for a detailed design specification.

**Figure 3.**
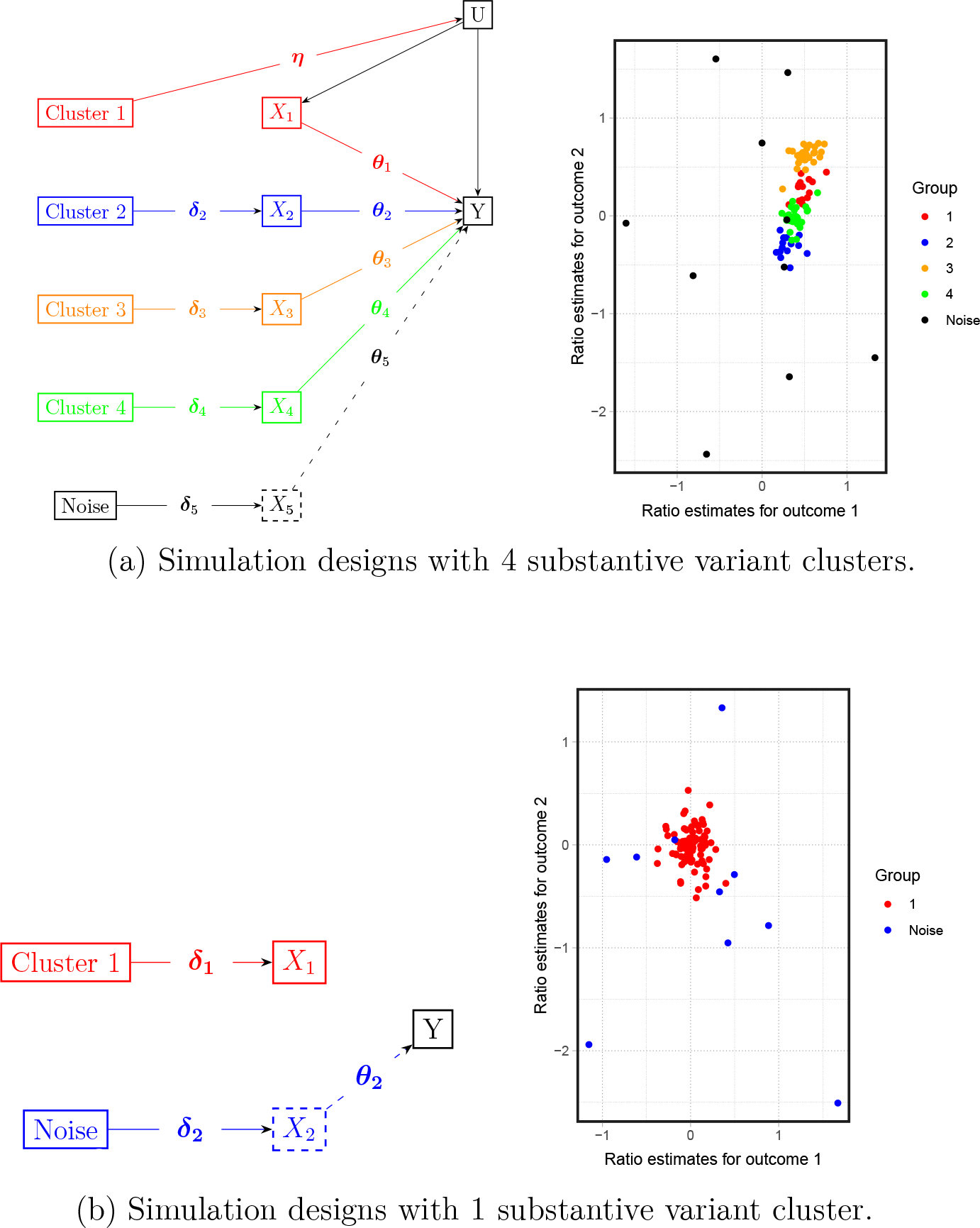
Directed acyclic graphs (DAG) of the data generation process and scatter plots of representative simulated data (with two outcomes) of the simulation designs with different numbers of variant clusters.

We compare MR-AHC with two other multi-dimensional clustering approaches. The first is the mclust algorithm [26], a general clustering method that accommodates the utilization of ratio estimates as inputs. We select this method as it is one of the most widely used clustering method based on Gaussian mixture models [26]. We implement mclust in two ways: the basic setting without a noise component, and the setting incorporating a Poisson noise component. An initial value of the proportion of noise variants is required, and we set this value favourably as 10% which is the ground truth. The second method, NAvMix, proposed by Grant et al. [25], groups genetic variants based on the variant-trait associations instead of the ratio estimates. We choose this method for comparison as it is also motivated by elucidating the biological mechanisms that can possibly be revealed by the patterns of the genetic variants associated with various traits. We employed two sets of input data for NAvMix: the variant-trait associations, as initially proposed; and the ratio estimates as in MR-AHC. Similar to mclust, we set the initial proportion of noise variants for NAvMix at 10%. For these two methods, cluster membership is assigned based on the highest probability. As a general clustering method for mechanistic heterogeneity, MR-AHC also works in the one-outcome case. We compare MR-AHC with MR-Clust [15], a popular method for conducting one-dimensional clustering based on ratio estimates, see Table S4 in Appendix D.

We report the following statistics from the simulations for each approach: the number of substantive clusters detected by the methods (“*#clusters*”); the Rand index which measures the similarity between the true clustering structure and the detected clusters for the substantives clusters (“*Rand index*”); the number of variants classified into the junk cluster (“*#junk variants*”); the number of true noise variants classified by the methods as junk (“*correct junk*”); the mean absolute error (“*MAE* “) and the mean squared error (“*MSE* “). For simulation designs with one substantive cluster indicating zero causal effects, we additionally report the frequency of correctly identifying the null cluster (“*Freq*.*null* “). Definitions of the statistics can be found in the method section.

For scenarios involving non-zero outcome sample correlations, since the GWAS results typically lack a direct estimate for the correlation, we initially apply all methods treating the correlation *ρ* as zero. The two-outcome simulation results are presented in Figure 4 and Table 1. Across all settings, MR-AHC consistently demonstrates high accuracy in identifying the number of clusters, aligning closely with the ground truth in both mean and median assessments. In the boxplot of MR-AHC (Panel (a) of Figure 4), all the quantiles are concentrated around the median, showing that the method consistently divides variants into the correct number of clusters with little fluctuation. By comparison, both settings of mclust tend to underestimate the number of clusters when there are four true clusters and overestimate it when only one substantive cluster exists. The NAvMix method, employing two different sets of input, also exhibits a tendency to underestimate the number of clusters when *K* = 4. While it successfully identifies one cluster when *K* = 1 with low outcome correlations, it overestimates the cluster number when the out-come correlation is high (*ρ* = 0.7). In line with the cluster number results, MR-AHC performs very well in terms of grouping the variants correctly, as measured by the Rand index. It consistently achieves Rand indices close to 1, significantly outperforming all other approaches in all settings. One potential drawback of MR-AHC is its tendency to assign slightly more noise variants to the “junk” cluster than the true count, but the number of true noise variants selected as junk of MR-AHC is only marginally lower than that of mclust with the noise component, outperforming all other approaches. Regarding estimation bias, both MR-AHC and NAvMix with ratio estimates input exhibit comparable MAE and MSE in general, both of which are smaller than those of other methods in most of the settings. For scenarios where *K* = 1, MR-AHC accurately identifies the null cluster with frequencies close to 1. The two variations of NAvMix also exhibit high accuracy in this aspect, although this accuracy diminishes for NAvMix with ratio estimates when the outcome correlation is high.

**Table 1:** Simulation results for designs with two outcomes. All methods are conducted treating the outcome correlations as 0. “*mclust noise*” stands for the mclust algorithm with a noise component, and “*NAvMix ratio*” for the NAvMix method with ratio estimates as input. Statistics are calculated as the mean over 1000 replications.

|  | MR-AHC | mclust | mclust noise | NAvMix | NAvMix ratio |
| --- | --- | --- | --- | --- | --- |
| **Two outcomes, K = 4, correlation = 0** |  |  |  |  |  |
| # clusters | 4.196 | 3.716 | 2.964 | 3.018 | 2.307 |
| Rand index | 0.917 | 0.757 | 0.737 | 0.615 | 0.710 |
| # junk variants | 10.966 | 0.000 | 7.726 | 6.517 | 4.080 |
| # correct junk | 6.975 | 0.000 | 7.134 | 1.446 | 3.951 |
| MAE | 0.088 | 0.120 | 0.123 | 0.162 | 0.106 |
| MSE | 0.030 | 0.041 | 0.041 | 0.052 | 0.035 |
| **Two outcomes, K = 4, correlation = 0.2** |  |  |  |  |  |
| # clusters | 4.207 | 3.665 | 2.933 | 3.130 | 2.254 |
| Rand index | 0.915 | 0.744 | 0.725 | 0.630 | 0.694 |
| # junk variants | 10.916 | 0.000 | 7.835 | 6.930 | 3.999 |
| # correct junk | 7.031 | 0.000 | 7.234 | 1.493 | 3.906 |
| MAE | 0.088 | 0.118 | 0.122 | 0.153 | 0.101 |
| MSE | 0.030 | 0.039 | 0.040 | 0.048 | 0.032 |
| **Two outcomes, K = 4, correlation = 0.7** |  |  |  |  |  |
| # clusters | 4.229 | 3.652 | 3.754 | 3.649 | 2.202 |
| Rand index | 0.918 | 0.784 | 0.835 | 0.684 | 0.677 |
| # junk variants | 10.993 | 0.000 | 8.044 | 5.347 | 3.925 |
| # correct junk | 7.144 | 0.000 | 7.433 | 1.302 | 3.844 |
| MAE | 0.089 | 0.091 | 0.088 | 0.119 | 0.087 |
| MSE | 0.029 | 0.027 | 0.027 | 0.035 | 0.024 |
| **Two outcomes, K = 1, correlation = 0** |  |  |  |  |  |
| # clusters | 1.026 | 2.211 | 1.312 | 1.000 | 1.001 |
| Rand index | 0.947 | 0.887 | 0.895 | 0.818 | 0.818 |
| # junk variants | 10.194 | 0.000 | 8.211 | 0.000 | 0.000 |
| # correct junk | 7.703 | 0.000 | 7.073 | 0.000 | 0.000 |
| MAE | 0.012 | 0.020 | 0.015 | 0.017 | 0.017 |
| MSE | 0.000 | 0.004 | 0.001 | 0.000 | 0.000 |
| Freq.null | 0.955 | 0.726 | 0.767 | 0.998 | 0.997 |
| **Two outcomes, K = 1, correlation = 0.2** |  |  |  |  |  |
| # clusters | 1.019 | 2.226 | 1.297 | 1.009 | 1.007 |
| Rand index | 0.949 | 0.881 | 0.893 | 0.816 | 0.816 |
| # junk variants | 10.275 | 0.000 | 8.438 | 0.058 | 0.060 |
| # correct junk | 7.833 | 0.000 | 7.199 | 0.009 | 0.008 |
| MAE | 0.012 | 0.021 | 0.015 | 0.017 | 0.018 |
| MSE | 0.000 | 0.004 | 0.001 | 0.000 | 0.001 |
| Freq.null | 0.965 | 0.724 | 0.791 | 0.996 | 0.990 |
| **Two outcomes, K = 1, correlation = 0.7** |  |  |  |  |  |
| # clusters | 1.025 | 2.221 | 1.296 | 1.883 | 1.881 |
| Rand index | 0.948 | 0.910 | 0.924 | 0.546 | 0.545 |
| # junk variants | 9.685 | 0.000 | 9.458 | 29.057 | 27.936 |
| # correct junk | 7.995 | 0.000 | 8.323 | 5.099 | 5.020 |
| MAE | 0.013 | 0.019 | 0.014 | 0.026 | 0.113 |
| MSE | 0.000 | 0.003 | 0.001 | 0.001 | 0.015 |
| Freq.null | 0.954 | 0.760 | 0.785 | 0.928 | 0.102 |

**Figure 4.**
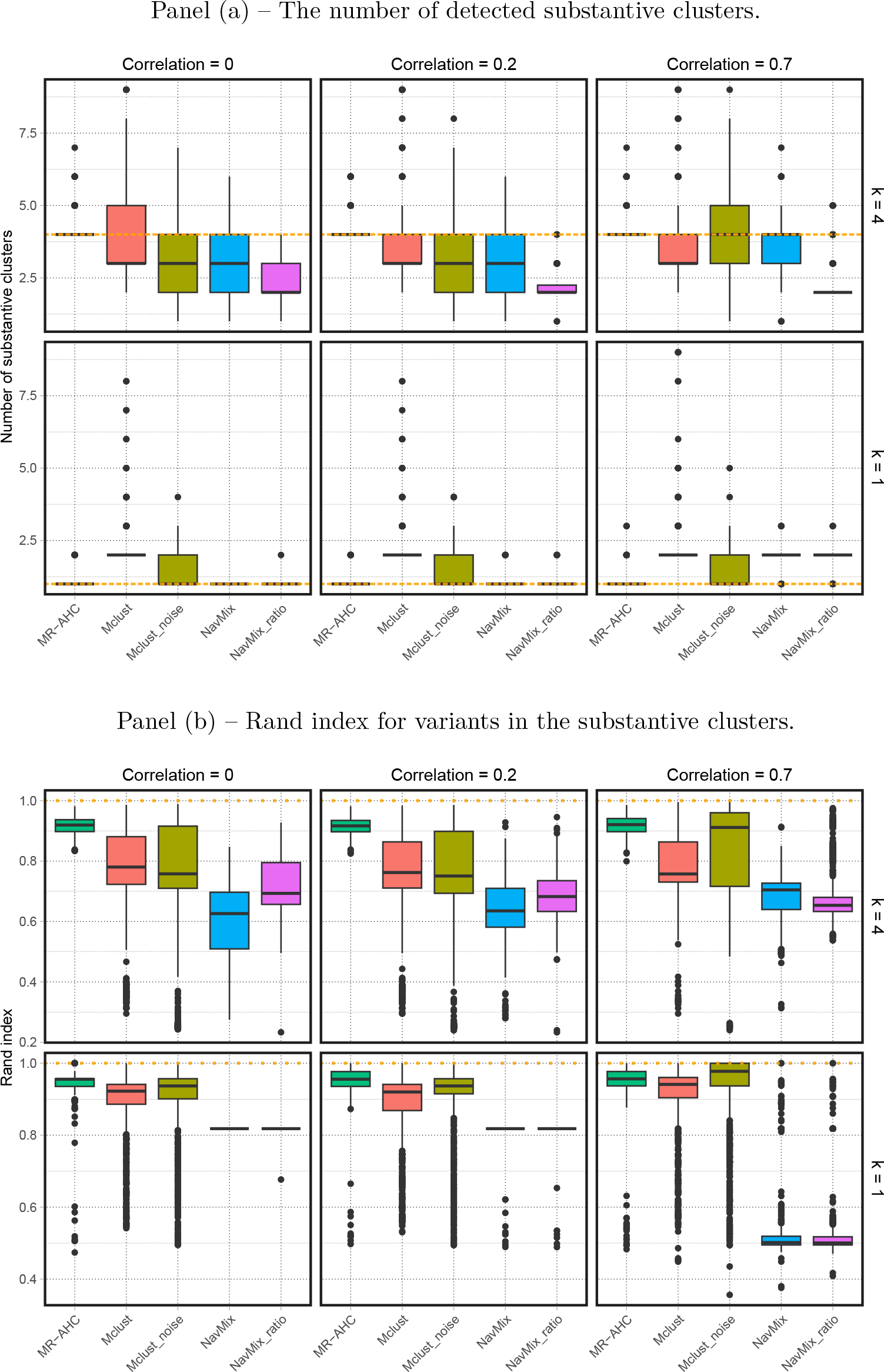
Two outcomes – boxplots for the number of detected substantive clusters and Rand index with different cluster numbers (*K* = 4 or *K* = 1) and outcome correlations (*ρ* = 0, *ρ* = 0.2 and *ρ* = 0.7). All methods are conducted treating the outcome correlations as 0. The dotted horizontal lines represent the true values. “*mclust noise*” stands for the mclust algorithm with a noise component, and “*NAvMix ratio*” for the NAvMix method with ratio estimates as input. Results are based on 1000 replications.

When *K* = 4 with non-zero outcome correlations, MR-AHC tends to identify more clusters than the ground truth. This feature can be rectified by incorporating accurate outcome correlation information, see the results generated by applying the method with the true correlation parameter (Table S2 in Appendix D). We show in Appendix A that the outcome correlation depends on both the extent of sample overlap between the outcome samples, and the phenotypic correlation between the outcome traits. Hence, high outcome correlations are uncommon in practice. To achieve, for instance, a correlation of *ρ* = 0.7, one would need perfect sample overlap and a phenotypic correlation of 0.7 between the two outcome traits. Even in this extreme scenario, implementing MR-AHC while assuming a zero correlation performs reasonably well. The simulation results for scenarios with three outcomes are presented in Appendix D, Table S1 and S3. Once again, MR-AHC exhibits good performance, producing clustering results that closely align with the ground truth and generally surpassing the performance of all other approaches.

### Estimating the causal effects of higher adiposity on type 2 diabetes and osteoarthritis

We apply the MR-AHC method to investigate the causal relationship between body fat percentage (BFP), as a measure of adiposity, and a pair of multimorbid conditions, T2D and OA. We use a three-sample summary-data MR design with 487 SNPs associated with BFP as instruments, accounting for the causal effects of the common risk factor BFP on both of the conditions simultaneously. For comparison, we also perform variant clustering using the mclust algorithm and the NAvMix method.

The clustering results of MR-AHC are presented in Figure 5, Panel (a). It detects 4 substantive clusters indicating heterogeneous causal effects. The cluster-specific estimation results, obtained with the inverse-variance weighted (IVW) approach [5], are depicted in Figure 5, Panel (b). Among the 4 clusters, Cluster 1 with 124 SNPs is the only cluster associated with increasing risk for both conditions; Cluster 2 with 258 SNPs indicates an increasing risk for T2D but a null effect for OA; both Cluster 3 (32 SNPs) and Cluster 4 (22 SNPs) are associated with a protective effect against T2D, and for OA, a causative effect and a null effect, respectively. See Appendix E for detailed estimation results.

**Figure 5.**
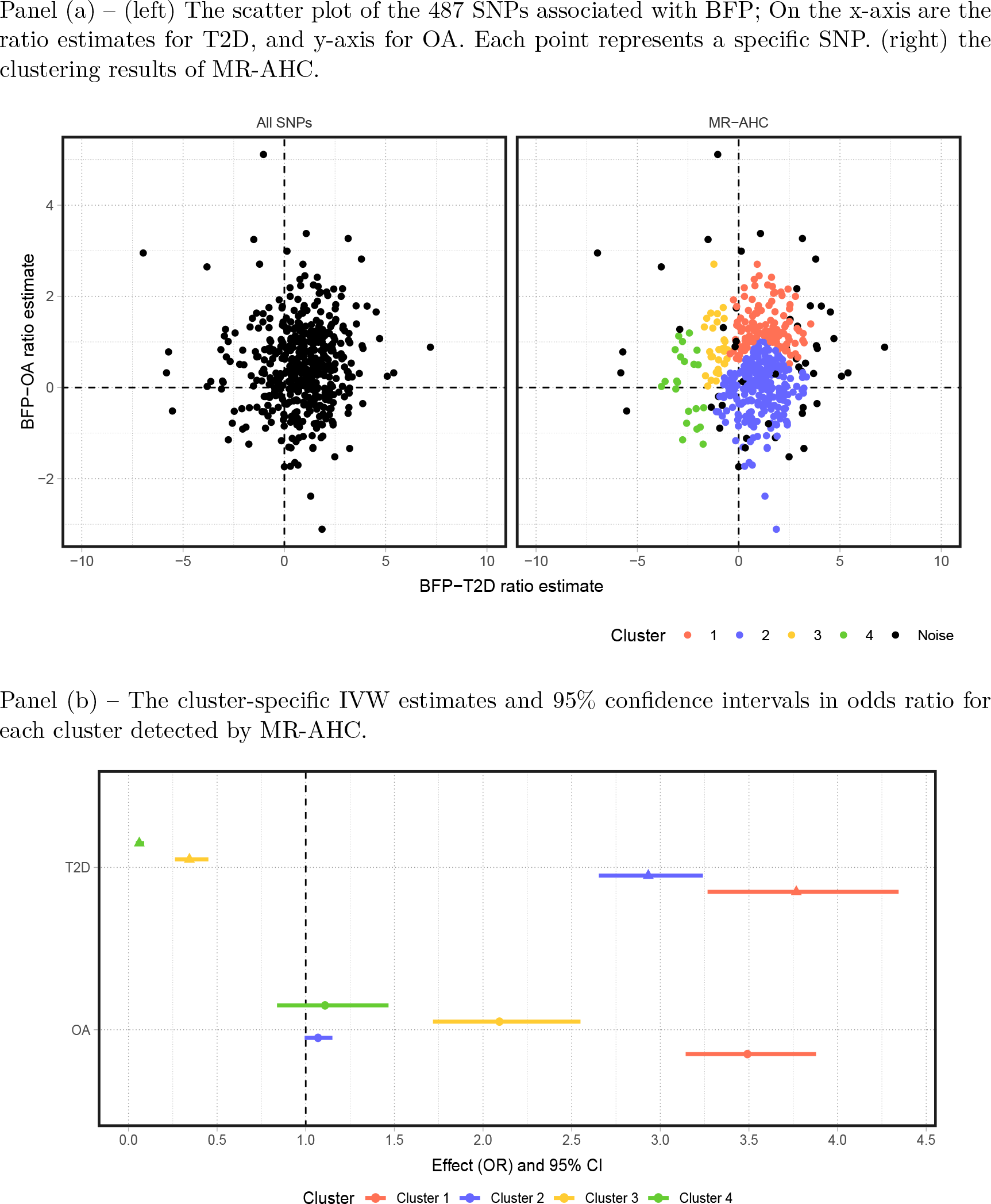
MR-AHC clustering and estimation results of the 487 SNPs associated with BFP based on their ratio estimates on T2D and OA.

These results align with the conclusions drawn from previous research. For example, Martin et al. [24] examined the causal effects of higher adiposity on a variety of conditions including T2D and OA. Their findings suggest that adiposity exerts heterogeneous effects on the risk of T2D: in general, higher adiposity increases the risk of T2D, but there is a metabolically “favourable” component of adiposity that reduces the risk of the condition. For OA, all adiposity measures, including the metabolically favourable adiposity, consistently identify an increasing risk. This suggests a non-metabolic weight-bearing effect as a likely cause. Given this, it is reasonable to partition the variants into distinct clusters along both outcome dimensions: on the T2D-estimate dimension, clustering occurs due to the indication of opposing effects by different variants; on the OA-estimate dimension, clustering is also likely to occur, as we may expect an adverse effect if the variants are associated with fat located around the articulations in a load-bearing way, but no effect elsewhere.

The clustering results generated with mclust and NAvMix are presented in Figure 6. Both methods fail to segregate the variants along the OA-estimate dimension, as all clusters indicate increasing effects, hence might have underestimated the number of clusters, which also appears as an over-arching feature of the methods in the simulations. Even for the T2D-estimate clustering, their results may be dubious: mclust assigns SNPs in nonadjacent regions with largely opposing estimates into the same cluster (Cluster 2 in blue); NAvMix either labels a large number of SNPs as ‘junk’ if setting a non-zero initial noise proportion, or does not identify any noise at all with a zero initial proportion. More importantly, for clusters generated by these two methods, variants tend to display substantial within-cluster heterogeneity in their ratio estimates, which can be a significant concern for causal inference.

**Figure 6.**
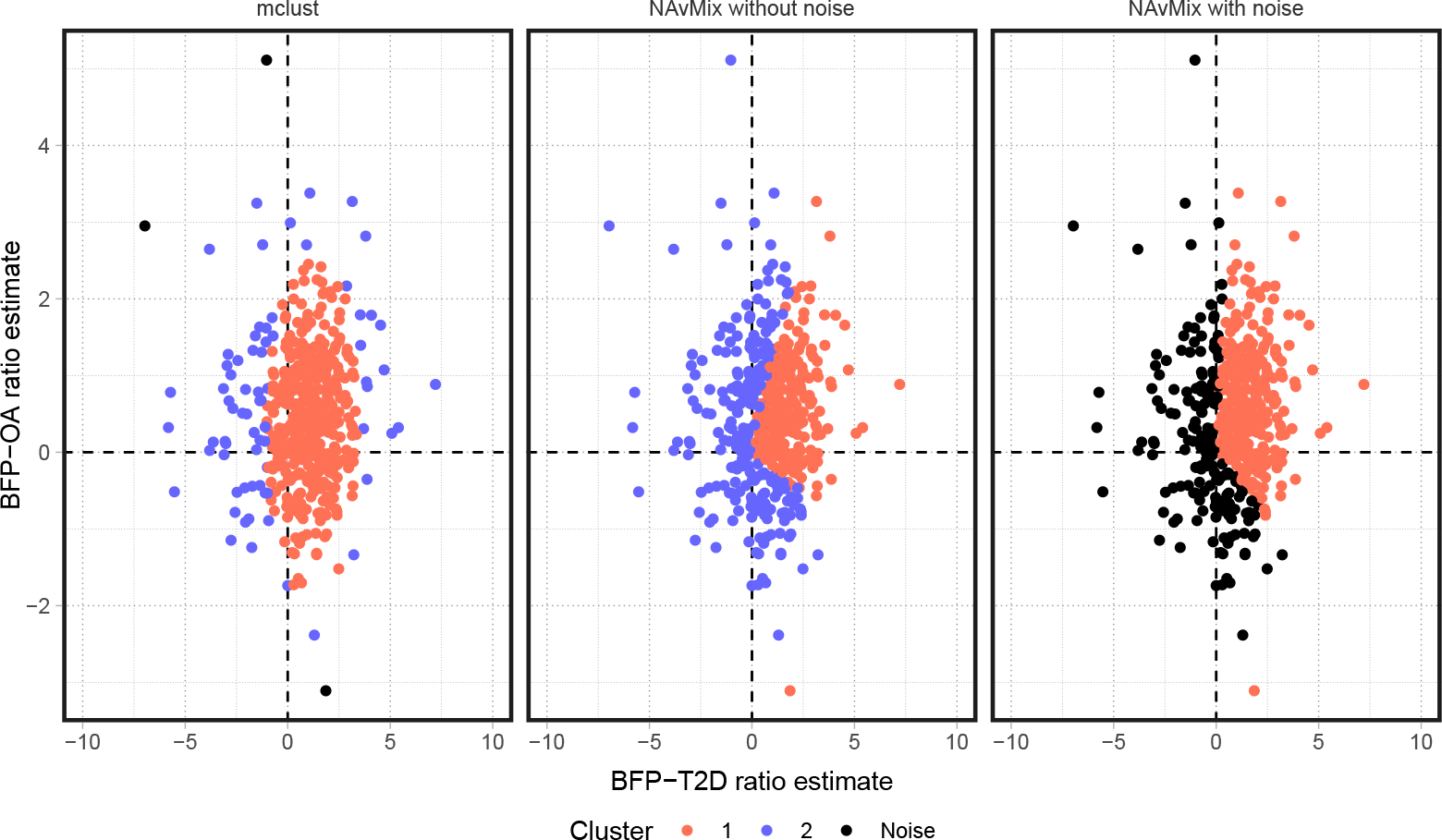
From left to right: the clustering results of the mclust algorithm with an initial noise proportion 5%; the clustering results of the NAvMix method with an initial noise proportion 0; the clustering results of the NAvMix method with an initial noise proportion 5%.

### Biological insights into the variant clusters

To gain insights into the biological mechanisms linking obesity to the T2D-OA multimorbidity from the variant clusters detected by MR-AHC, we use an approach similar to the one taken by previous works such as Grant et al. [25] and Wang et al. [31]. For each of the clusters identified by MR-AHC, we first map the SNPs in the cluster to genes, then perform gene set enrichment analysis with the mapped genes. Both steps are conducted using the Functional Mapping and Annotation Platform (FUMA) [32]. SNPs are mapped to genes using a three-way mapping strategy (positional, eQTL and chromatin interactions mapping). The gene set enrichment analysis is to test if the mapped genes are over-represented in a given pre-defined gene set which corresponds to a canonical biological pathway or is associated with a phenotype reported from the GWAS catalog. We refer to the latter as the gene-set Phenome-wide association analysis (PheWAS), or just “PheWAS” for short. We integrate both lines of evidence from the pathway and PheWAS analyses that can complement or validate each other, to infer the possible biological mechanisms underlying each cluster. See Supplementary material S2 for a summary of the enrichment analyses results.

First, it is likely that Cluster 2 (containing 258 SNPs, associated with increasing risk of T2D) is highly pleiotropic. Based on the PheWAS analysis, this cluster is enriched with a large number of phenotypes, double that for Cluster 1 which has the second most (112 versus 56). These phenotypes fall into a wide range of categories, displaying no clear pattern. The majority of the canonical pathways enriched for this cluster are related to intermediate filament, which might not have a strong direct link with the causal relationship under examination.

Cluster 1 (containing 124 SNPs, indicating increasing risks of both conditions) holds particular significance as it aligns with our primary objective of exploring the multimorbidity of T2D and OA through obesity. The majority of the canonical pathways uniquely enriched for Cluster 1 can be classified into two categories of cellular processes that are closely interconnected: gene expression transcription and cellular responses to stimuli. A significant example in the first category is DNA methylation, while in the second category, one of the most significantly enriched pathways is associated with oxidative stress. For some of the pathways, we can delve deeper into the investigation using readily available GWAS data. As an example, we further inspect the possible unifying pathway from obesity to the T2D-OA multimorbidity via oxidative stress.

Oxidative stress (OS) is the imbalance between the production of reactive oxygen species and the counteracting antioxidant defenses in the direction that favors the former, which may lead to tissue injury [33]. Clinical research has established that obesity can induce systemic OS through various metabolic pathways [34, 35]. Moreover, OS is evidenced to exert direct effects on the development of T2D via mechanisms such as decreasing insulin secretion from pancreatic *β* cells [36, 37]. It also plays a role in the progression of OA by promoting cartilage degradation [38]. Herein, we examine the role of OS by performing cluster-specific MR: we first analyze how BFP predicted by SNPs in cluster 1 is associated with a variety of OS biomarkers. Then for comparison, we conduct the same analysis on Cluster 4, serving as a counterpart to Cluster 1 due to its relatively benign nature for both conditions, manifesting a protective effect against T2D and a null effect on OA.

We select 11 OS biomarkers from diverse categories. First, as endogenous antioxidants are highly responsive to OS [39], we use 4 enzyme antioxidants (GST, CAT, SOD, GPX) as OS injury biomarkers, which have been utilized in previous MR studies [40, 41]. One of the mechanisms through which obesity induces systemic OS is chronic inflammation [34, 35]. We thus incorporate three traits known to mediate the pathway from inflammation to OS (CRP, IL-6, TNF-*α*) [35] as another set of OS biomarkers. Biochemical research has shown that the production of some cytokines, including IL-1*β*, IL-12 and IL-8, are enhanced under elevated OS levels [42, 43]. Therefore, we also include these three cytokines in the analysis. Finally, we incorporate GDF-15, which is a biomarker for both inflammation and OS [44]. See the method section for the full form of the abbreviations of the biomarkers.

We estimate the effect of BFP on each of the biomarkers by two-sample MR using SNPs in Cluster 1 and Cluster 4 as instruments separately. The estimates and standard errors are calculated by the IVW approach. Sensitivity checks by MR-PRESSO [45] and the robust adjusted profile score (MR-RAPS) method [13] can be found in Appendix E. Results in Z-scores are presented in Figure 7. For 8 out of 11 of these OS markers, Cluster 1 is associated with increasing effects, while Cluster 4 is associated with declining effects. For CAT and CRP, Cluster 1 and Cluster 4 have effects in the same direction, but Cluster 1 is either associated with a larger increasing effect (CRP), or a smaller decreasing effect (CAT). The only exception is SOD, on which the increasing effect of Cluster 1 is smaller than that of Cluster 4.

**Figure 7.**
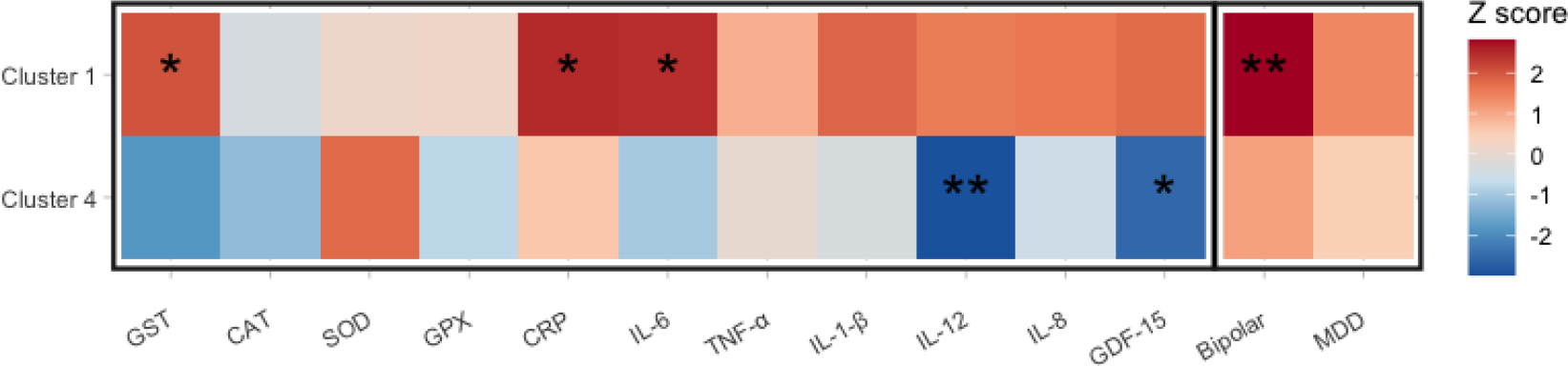
Results of the two-sample MR estimating the effects of BFP on the 11 oxidative stress biomarkers and 2 psychological disorders using variants in Cluster 1 and Cluster 4 as instruments respectively. Estimates are given by the IVW approach, presented in the form of Z-scores (the ratio of the estimate and the standard error). “*” represents significance at the p-value 0.05; “**” for the first 11 traits represents significance at 0.05*/*11, for the last two traits at 0.05*/*2.

Overall, we can see a clear heterogeneity pattern between Cluster 1 and Cluster 4 in their cluster-specific effects on the OS biomarkers, which supports that Cluster 1 is associated with an elevated level of oxidative stress, while it may be the opposite for Cluster These results align with the existing findings regarding adiposity and oxidative stress: higher adiposity is in general associated with elevated oxidative stress, but fat patterns featured with a smaller waist-to-hip ratio (WHR) may be related to less oxidative damage [35, 46]. This correlation between WHR and OS is observed in Cluster 4, as we will show later that this cluster is associated with a decreasing WHR.

Complementary evidence that may be related to the shared pathway via oxidative stress can be found in the PheWAS results for Cluster 1. A notable PheWAS pattern associated with this cluster is that it is enriched with quite a few psychological disorders. Clinical research has shown that OS is implicated in the development of such disorders, including bipolar disorder and depression [47], which are both significantly enriched for Cluster 1. We estimate the effects of BFP predicted by variants in respectively Cluster 1 and Cluster 4 on bipolar and major depressive disorder (MDD) using two-sample MR. Results are presented in the last two columns in Figure 7. Cluster 1 is associated with increasing risks of both conditions with a significant effect on bipolar. The effects of Cluster 4, on the other hand, are both insignificant and smaller than those of Cluster 1. These results may suggest a possible direction for exploring the multimorbidity between obesity-related metabolic conditions and psychological disorders.

It is important to note that there is very likely to be intricate interactions between the pathways involved in the underlying mechanism from obesity to the T2D-OA multimorbidity. For example, another canonical pathway uniquely enriched for Cluster 1 is related to programmed cell death, or apoptosis. It has been well-documented that excess OS plays a role in the activation of apoptosis [48], and pancreatic *β*-cell and chondrocyte loss due to apoptosis are implicated in the development of T2D and OA respectively [49, 50]. Furthermore, quite a few gene expression transcription pathways enriched for Cluster 1 are related to epigenetic processes. Emerging evidence supports the involvement of OS in epigenetic regulation of gene expression such as inducing DNA methylation changes [51, 52]. Thus, additional research is warranted to further unravel the exact causal roles of these pathways.

Both Cluster 3 and Cluster 4 exhibit a protective effect against T2D. The most noteworthy PheWAS pattern for these two clusters is that they are both enriched with phenotypes related to fat distribution. This is particularly pronounced for Cluster 4, with 17 out of 42 enriched phenotypes associated with fat patterns including the WHR-related traits. Also, Cluster 4 has a clear pattern regarding its enriched biological pathways: 13 out of 16 of the pathways are related to ion channel activities. Ion channels are membrane proteins acting as gated pathways for the passage of ions across the cell membranes [53].

To integrate the evidence from the WHR-enriched PheWAS pattern and the ion-channel-enriched pathway pattern into a potential explanation of the protective mechanism against T2D, one possible link may be that Cluster 4 is also enriched with several HDL-C related phenotypes. Existing studies have found a negative relationship between WHR and HDL-C [54, 55], i.e. smaller WHR may be associated with higher levels of HDL-C. Moreover, the connection between HDL-C levels, ion channel activities, and T2D development might be explained by the primary role of HDL-C in cholesterol clearance [56]. On one hand, ion channels, such as the *β*-cell voltage-gated calcium channels, are crucial for insulin secretion [57]. On the other hand, the activity of such channels can be suppressed by excess membrane cholesterol [29]. Thus, the depletion of cholesterol facilitated by HDL-C might positively impact the activity of the ion channels related to insulin secretion. This link is evidenced by previous experimental research on mice, which shows that reduced HDL-C levels are correlated with impaired glucose-induced insulin secretion [58]. This is because the increased rigidity of the *β*-cell membrane due to cholesterol-enrichment reduces the stimulation of ion channels essential for secreting insulin [59, 60].

To examine the possible protective mechanism against T2D stated above, we conduct two-sample MR to examine the effects indicated by Cluster 4 on WHR (adjusted for BMI), HDL-C and total cholesterol levels. The results, shown in Figure 8, are in line with the hypothesized mechanism: this cluster is associated smaller WHRs, higher levels of HDL-C, lower levels of total cholesterol, and consequently decreasing risk of coronary artery disease (CAD).

**Figure 8.**
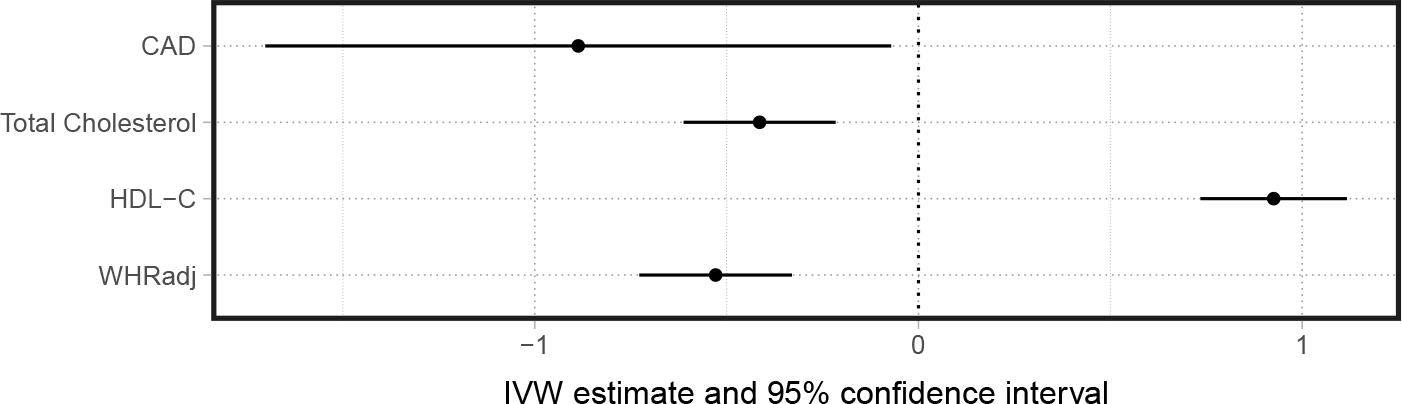
Two-sample MR results estimating the effects of cluster 4-predicted BFP on the waist-to-hip ratio adjusted for BMI, HDL-C, total cholesterol and coronary artery disease. Estimates are given by the IVW approach, presented in the form of the 95% confidence intervals.

## Discussion

In this paper we adapt the general method of agglomerative hierarchical clustering to the summary-data Mendelian randomization setup. MR-AHC is a useful tool for interrogating a set of genetic variants to see if they collectively identify a single causal effect, or if it is more plausible that a number of subgroups identify distinct effects driven by different biological mechanisms. The method is of particular interest when the potentially heterogeneous physiological components of the exposure are not known beforehand, or are difficult/expensive to measure. Of special interest is its utility as a multi-dimensional clustering method in the multi-outcome MR setting, where we can elucidate the shared causal pathways that underlie the co-occurrence of a range of conditions through a common risk factor.

In an effort to investigate the intricate mechanisms underpinning disease causation, a number of approaches has been utilized to categorize genetic variants associated with a specific phenotype, based on their GWAS associations with a range of traits linked to that target phenotype, such as the Bayesian nonnegative matrix factorization clustering method [61] and the NAvMix method [25]. MR-AHC is motivated similarly by the purpose of exploring the diverse disease-causing pathways reflected by distinct variant clusters. However, it is distinctly tailored to a different scenario and employs a different clustering strategy. Its primary application is rooted in the domain of causal inference, specifically within the framework of MR. It groups genetic variants based on their causal estimates, which integrates both their associations with the target phenotype (in this context, a common exposure) and their associations with the related traits (herein, downstream outcomes). By the comparison with NAvMix through Monte Carlo simulations and the real-world application, we have shown that MR-AHC has certain advantages over the association-based approaches in MR settings, namely an enhanced capacity to identify the patterns of the genetic variants that may mirror distinct causal mechanisms between specific traits. Furthermore, while the variant clusters discovered through association-based methods may have broad biological implications encompassing a wide range of traits, those detected by MR-AHC are precisely focused on elucidating a specific causal relationship. Consequently, pathway information derived from each cluster identified by MR-AHC offers a higher degree of relevance and specificity for the causal relationship under examination.

MR-AHC possesses the features that it does not require pre-specifying the number of clusters, and that alongside detecting meaningful clusters it can also identify and label null and junk clusters without an initial specification on the proportion of ‘noise’. While for hierarchical clustering algorithms, it can be difficult to choose the ‘optimal’ dissimilarity metric, linkage and number of clusters on the dendrogram to yield reliable clustering results, studies in the field of model selection [17, 62] provide the theoretical basis for MR-AHC to ensure highly accurate results. We have adapted the original AHC method in Apfel and Liang [17] to accommodate the varying degrees of uncertainty exhibited in summary-data estimates due to allele frequency differences across SNPs. Moreover, our method is capable of handling outliers in the variant-specific estimates with our outlier removal procedure. It should be noted that all the aforementioned methods assign variants to clusters in a probabilistic (i.e. ‘soft’) way, while MR-AHC do the clustering in a deterministic (i.e. ‘hard’) manner. Although we view this as a strength, some may view its lack of stochasticity as a disadvantage. For this reason we plan to develop a framework to quantify the sensitivity of MR-AHC clustering results to small changes in the data and thresholding rule used.

We showed in simulations that, in situations of sample overlap in the outcome data, incorporating the correct correlation information can improve the performance of the method. Nevertheless, it is in general not a significant concern if the correlation estimates are set to zero. Our method is currently focused on the problem of estimating a causal relationship between the shared exposure and the downstream outcomes without accounting for the direct causality between the outcomes. In our application example, various existing evidence supports the absence of direct causality between T2D and OA [63, 64]. However, we show in Appendix C that even if direct causality exists, our method is still applicable, as the clustering of the variants associated with the common exposure are generally robust to the outcome causality. The challenge then shifts to estimating the direct causal effect of the exposure on a particular outcome while considering other outcome traits as an additional risk factor, or accepting that the original estimates represent total causal effects via the outcome in question. Given this, another potential future extension of our work is to extend the method to the multi-exposure framework, with the additional flexibility to consider genetic sub-structure within each exposure.

## Materials and Methods

### Model setup

We start from the individual-level model underlying the variant-exposure and variant-outcome summary associations. We assume a general linear IV model for the MR framework allowing for multiple outcomes with a shared exposure, which accounts for pleiotropy and heterogeneous causal mechanisms. We also assume that the causal effects from the exposure to the outcomes via different pathways are additive. Therefore, without loss of generality, we model the causality heterogeneity using additive sub-components of the exposure. Let the common exposure *X* be denoted by *X* = *X*_1_ + … + *X*_*k*_ where *X*_*k*_ is the *k*-th sub-component in *X* and *k* = 1, …, *K*. Let the *p*-th observed outcome be denoted by the scalar *Y*_*p*_ where *p* = 1, …, *P* and *P ≥* 2 in a multi-outcome MR. The vector **G** = (*G*_1_, …., *G*_*J*_)^*′*^ is used to denote the *J* genetic variants used as instruments. We then have the following linear structural model:

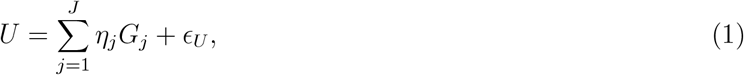

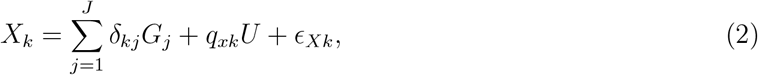

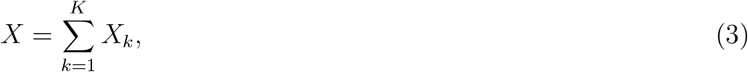

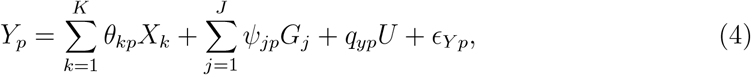

where

- U represents the uncontrolled confounding between *Y*_*p*_ and any sub-component of *X*, with the strength of confounding determined by parameters *q*_*xk*_ and *q*_*yp*_;
- *ϵ*_*U*_, *ϵ*_*Xk*_ and *ϵ*_*Y p*_ are error terms affecting *U, X*_*k*_ and *Y* respectively, and we assume *E*(*ϵ*_*U*_ *G*_*j*_) = *E*(*ϵ*_*Xk*_ *G*_*j*_) = 0 for all *j* = 1, …*J*;
- *θ*_1*p*_, …, *θ*_*Kp*_ represent the heterogeneous causal effects of *X* on *Y*_*p*_.

We maintain the assumption that all the variants are independent with each other, and therefore inspect the relationship between each individual variant *G*_*j*_ and *X* as well as between *G*_*j*_ and *Y*_*p*_. First, from (1) and (2), we obtain the reduced-form relationship between *X*_*k*_ and *G*_*j*_ as

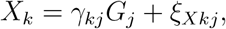

where the total effect of *G*_*j*_ on *X*_*k*_ is *γ*_*kj*_ = *δ*_*kj*_ + *q*_*xk*_ *η*_*j*_. The error term *ξ*_*Xkj*_ is defined implicitly, but from the the previous assumptions *E*(*ϵ*_*U*_ *G*_*j*_) = *E*(*ϵ*_*Xk*_*G*_*j*_) = 0 and that *G*_*j*_ is independent with all other variants, we have *E*(*ξ*_*Xkj*_ *G*_*j*_) = 0 as well. It follows from (3) that the overall relationship between *G*_*j*_ and the exposure *X* is

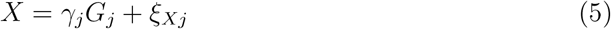

where

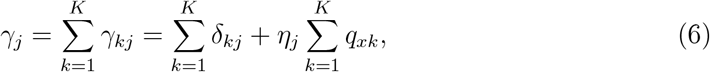

and the error term 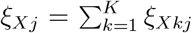with *E*(*ξ*_*Xj*_ *G*_*j*_) = 0. We assume that the relevance condition for the instruments is satisfied at the scale of the overall exposure *X* so that *γ*_*j*_*/*= 0 for *j* = 1, …, *J*.

Now we inspect the reduced-relationship between *G*_*j*_ and *Y*_*p*_. It follows from (4) that the pleiotropic effect of *G*_*j*_ on *Y*_*p*_ can be derived as *α*_*jp*_ = *Ψ*_*jp*_ + *q*_*yp*_*η*_*j*_. Additionally by plugging (1) and (2) into (4), the overall reduced-form between *Y*_*p*_ and *G*_*j*_ is

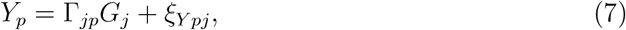

Where

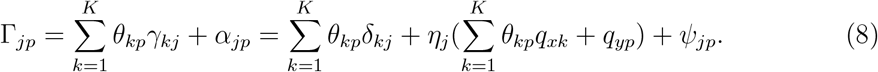

The correlation between the implicitly-defined error term *ξ*_*Y pj*_ and *G*_*j*_ depends on the correlation between *ϵ*_*Y p*_ from Equation 4 and *G*_*j*_. If *E*(*ϵ*_*Y p*_*G*_*j*_) = 0, then *ξ*_*Y pj*_ and *G*_*j*_ are also uncorrelated. In this case, for the *G*_*j*_ -*Y*_*p*_ association estimate (denoted by 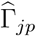) generated from a GWAS by regressing *Y*_*p*_ on *G*_*j*_ in a given sample, we have

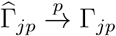

as the sample size *n → ∞*. Similarly, for the *G*_*j*_ -*X* association estimated in a GWAS (denoted by 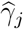) in a sample independent from the *G* -*Y* sample, we have

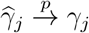

as *n → ∞*. Then for the variant-specific causal estimate of *G*_*j*_, defined as 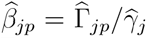, we have

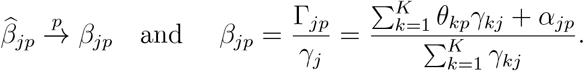

In words, as the sample size *n* goes to infinity, each 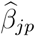 converges to their variant-specific causal estimand *β*_*jp*_, which is the causal effect from *X* to *Y*_*p*_ identified using *G*_*j*_ as instrument. In the simple case where *G*_*j*_ only instruments one sub-component *X*_*k*_, the variant-specific causal estimand then becomes:

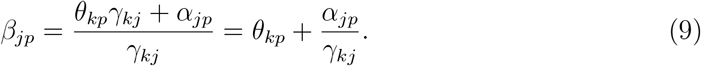

Equation 9 reflects the possible sources of mechanistic heterogeneity among the variant-specific estimates: heterogeneous causal effect from the exposure to the outcome, and pleiotropic effects. We aim to group the genetic variants into distinct clusters such that within each cluster, all variants identify the same causal effect. More generally, for the multi-outcome MR with *P ≥* 2 outcomes, with a given variant *G*_*j*_, we combine all 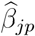 and *β*_*jp*_ for *p* = 1, …, *P* into the vectors 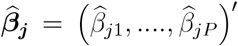 and ***β***_***j***_ = (*β*_*j*1_, …., *β*_*jP*_)^*′*^ respectively. We propose the MR-AHC method, elaborated in the subsequent section, to divide the genetic variants into distinct clusters based on the similarity of their ratio estimates 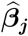, so that variants with the same estimand ***β***_***j***_ are in the same cluster.

Thus far, we have inspected the case where there is no residual correlation between *G*_*j*_ and *ϵ*_*Y p*_ in Equation 4, i.e. *E*(*ϵ*_*Y p*_*G*_*j*_) = 0. In a multi-outcome MR model, this relationship can be violated if there is direct causality between the outcome variables. For example, consider two outcomes *Y*_*p*_ and *Y*_*q*_, if *Y*_*q*_ causally affects *Y*_*p*_ directly, then it will enter Equation 4 as part of *ϵ*_*Y p*_, hence the error term may be correlated with *G*_*j*_. We show in Appendix C that the clustering results of the variants are in general not affected by the additional direct causality between the outcomes, but the causal effects identified by each cluster are the total effects including the outcomes causality, instead of the direct effects from the exposure to the outcomes. In this paper, we mainly focus on the case without the direct outcome causality.

### The MR-AHC algorithm

We make the following normality assumption on the summary statistics described in the previous section: 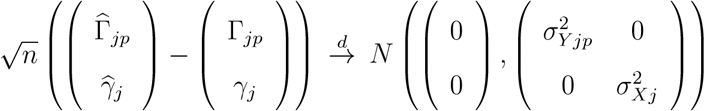,for *j* = 1, …, *J* and *p* = 1, …, *P*. It follows that 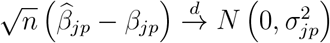. The estimates of the standard errors of 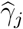 and 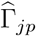, denoted by 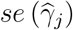 and 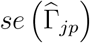, are generally given by the corresponding GWAS, hence taken as known. The standard error of 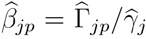 can then be obtained using the Delta method as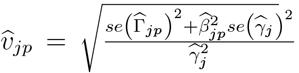 But it is typically approximated by 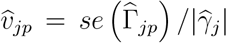 since 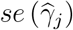 is deemed negligible when we only use variants that pass a genome-wide significance threshold as instruments [11]. When there are multiple outcomes, let 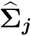 be the estimate of the covariance matrix of 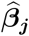. The diagonal entries of 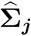 are just the variances of 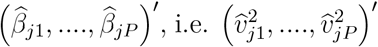 The off-diagonal entries of 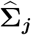 are the pairwise covariances of 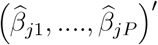. We show in Appendix A that the covariances are zero if the outcome samples are non-overlapping and/or the phenotypic correlations between the outcome traits are zero, otherwise, they can be estimated using LD score regression [28].

With the summary statistics 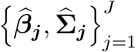 as inputs, we apply MR-AHC to discover the variant clusters using a two-step procedure: Step 1 (agglomerative hierarchical clustering) generates a decision path from *K* = *J* to *K* = 1 clusters; Step 2 (downward testing) re-traces the path from *K* = 1 to *K* = *J* until the optimal cluster choice *K*_*opt*_ is chosen. The first step is summarized as follows:

**Step 1**. *Ward’s algorithm [65]*

1. ***Initialization:*** *Each variant-specific estimate is viewed as a cluster on its own. Hence, initially, the total number of clusters is K* = *J*.
2. ***Merging:*** *The two clusters that are closest as measured by their weighted squared Euclidean distance are merged into a new cluster. Without loss of generality, assume this is satisfied by cluster 𝒮*_*k*_ *and* 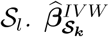*is defined as the inverse-variance weighted mean of all the variant-specific estimates in 𝒮*_*k*_, *as follows:*

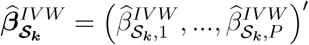

*where*

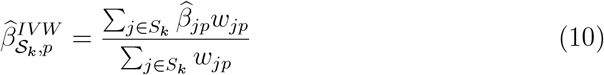

*with* 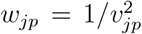 *for p* = 1, …, *P*. 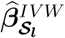*for Cluster 𝒮*_*l*_ *can be defined similarly. Then the weighted squared Euclidean distance between 𝒮*_*k*_ *and 𝒮*_*l*_ *is defined as*

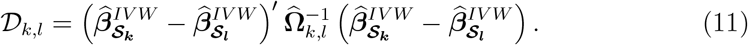

*The P × P matrix* 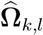*is defined as follows: let* 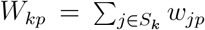, *and* 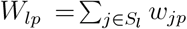 *for p* = 1, …, *P. Consider the entry at the i-th column and the r-th row of* 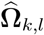*with i, r* ∈ {1, …, *P*}, *denoted by cov*_*ir*_. *We have*

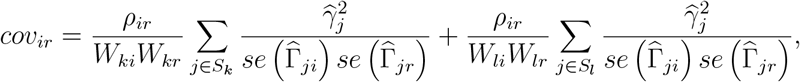

*where ρ*_*ir*_ *is the correlation between* 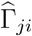 *and* 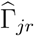, *which is assumed to be constant across j* = 1, …, *J. See Appendix A for details*.
3. ***Iteration:*** *The merging step is repeated until all the variant-specific estimates are in one cluster of size J*.

After generating the clustering path using Step 1, we are left with a *𝒦* = 1 super-cluster containing all variants. We then re-trace the pathway to select the optimal value of *K* using a downward testing procedure originally proposed by Andrews [62], operating as follows:

**Step 2**. *Downward testing procedure*.

*Firstly, define Q*_*fg*_ *to be the Cochran’s Q statistic [27] associated with the g-th cluster at level f of the clustering path, denoted by 𝒮*_*fg*_. *Also define T*_*fg*_ *to be the* (1 *−* ζ) *significance threshold of a χ*^2^ *distribution on P ×* (|*𝒮*_*f g*_ | *−* 1) *degrees of freedom with ζ*= 0.1*/log*(*n*), *n is the sample size, and* |*𝒮*_*f g*_ | *is the number of variants in 𝒮*_*fg*_. *Q*_*fg*_ *is defined as follows: for the p-th outcome, let* 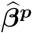*be the vector of length* |*𝒮*_*f g*_ | *with the j-th entry being* 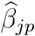 *where j ∈ 𝒮*_*fg*_. *Combine all* 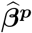*into a vector of length P ×* |*𝒮*_*f g*_ | *for all p* = 1, …, *P, denoted by ℬ*_*fg*_. *Let* 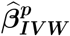 *be the IVW mean of all the estimates in* 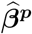*as defined in (10), and* ***ι*** *be a vector of* 1 *of length* |*𝒮*_*f g*_ |. *Then combine all the* |*𝒮*_*f g*_ |*-length vector* 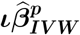 *into a vector of length P ×* |*𝒮*_*f g*_ | *for all p* = 1, …, *P, denoted by* 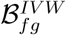. *Then Q*_*fg*_ *is*

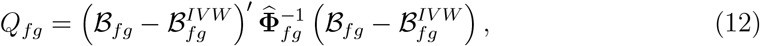

*where* 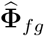*is a matrix that can be partitioned into P × P blocks. The block on the i-th column and r-th row, denoted by* 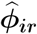, *is a* |*𝒮*_*f g*_ | *×* |*𝒮*_*f g*_ | *dimension diagonal matrix. The he diagonal entry equals to* 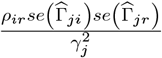 *for j* ∈ *𝒮*_*fg*_.

1. *Starting from the cluster that contains all the variants, calculate the global Q statistic, Q*_11_, *on all the ratio estimates;*
2. *If Q*_11_ *< T*_11_, *then stop and assume that all the variants form a single cluster. If Q*_11_ *≥ T*_11_, *then revert to the variant clusters on the next level of the path, where the number of clusters is 𝒦=2;*
3. *Calculate Q statistics for the two sub-clusters separately, Q*_21_ *and Q*_22_;
4. *If both Q*_21_ *< T*_21_ *and Q*_22_ *< T*_22_, *then stop. Otherwise, continue to the next level where 𝒦=3;*
5. *Repeat steps 3-4 until a 𝒦* ∈ (1, .., *J*) *is arrived at for which no sub-cluster heterogeneity statistic rejects at its given threshold*.

In implementing the MR-AHC method, in addition to the baseline procedure summarized in Step 1 and 2, we propose an extension of the method to handle outliers in the ratio estimates: after we run Step 1 and 2 and obtain the clustering results, within each detected cluster, calculate each individual variant’s contribution to the overall Q statistic. The individual Q statistic, calculated using (12) with only estimates of that variant, approximately follows a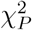distribution [11], and variants with large individual Q (here defined as the p-value of the individual Q below 5%) are viewed as outliers. We remove the outliers from each detected cluster, and re-run Step 1 and 2 with all the remaining variants. All the outliers are then assigned to the junk cluster.

### Monte Carlo simulations

For all the simulation designs, we simulate two/three-sample summary data based on the data generating process defined in Model (1)-(4) with all sample sizes equal to *N* = 60, 000. Here we fix *Ψ*_*j*_ = 0. We assume that *G*_*j*_, *X* and *Y*_*p*_ are normalized with *V ar*(*G*_*j*_) = *V ar*(*X*) = *V ar*(*Y*_*p*_) = 1 and 𝔼[*G*_*j*_] = 𝔼[*X*] = 𝔼[*Y*_*p*_] = 0. We also assume that the covariances between the variants are 0, *Cov*(*G*_*j*_, *G*_*i*_) = 0 for *i* ≠ *j*. According to Equation (6) and (8), the variant-exposure and variant-outcome summary statistics are generated in the following way:

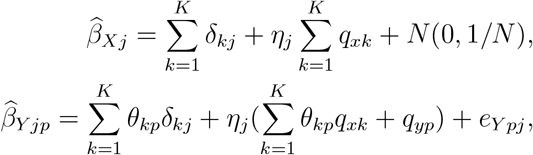

for *j* = 1, …., *J* with *J* = 100 and *p* = 1, 2 or *p* = 1, 2, 3. The normally distributed random variables *N* (0, 1*/N*) add the random component to 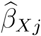 that mimics the asymptotically normal distribution of the statistics obtained from GWAS with standardized data. The random error of 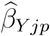 denoted by *e*_*Y pj*_, is generated from a multivariate normal distribution for the multiple outcomes. All the variance terms of this multivariate normal distribution are set to 1*/N*. When there are *P* = 2 outcomes, the covariance equals *ρ/N* with *ρ* = 0, 0.2, and 0.7 for the zero, low, and high outcome correlation settings respectively. When there are *P* = 3 outcomes, the pair-wise outcome correlation equals to *ρ*_*ij*_ = *ρ*^|*i−j*|^ where *i, j* ∈ {1, 2, 3} and *i* ≠ *j* with *ρ* = 0, 0.2 and 0.7 for three different settings. Then the standard errors of 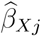 and 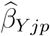are given by

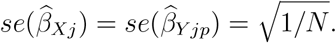

To simulate the summary statistics, we need to set the values of the following parameters: *θ*_*kp*_ (the causal effect of the sub-component *X*_*k*_ on *Y*_*p*_), *δ*_*kj*_ (the effect of variant *G*_*j*_ on *X*_*k*_), *η*_*j*_ (the effect of variant *G*_*j*_ on the uncontrolled confounder *U*), and *q*_*xk*_, *q*_*yp*_ (the effect of *U* on *X*_*k*_ and *Y*_*p*_ respectively). Let the variation in *U* explained by all the variants be 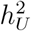, and the variation in *X*_*k*_ directly explained by all the variants (not through *U*) be 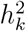. Then given the values of 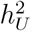 and 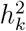, parameters *θ*_*kp*_, *q*_*xk*_ and *q*_*yp*_ are set as constants under the following restrictions to make *V ar*(*X*) = *V ar*(*Y*_*p*_) = 1 feasible:

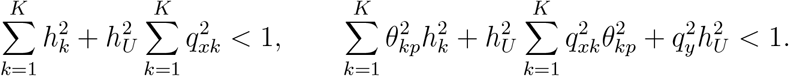

When there are *K* = 4 substantive clusters (with 15, 15, 30, 30 variants respectively) and one noise cluster (with 10 variants), let 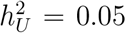 and 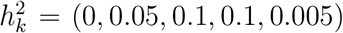 for *k* = 1, …, 5 with the last entry for the junk cluster. Set *q*_*xk*_ = 1 for the first cluster, which corresponds to the correlated pleiotropy pathway, and *q*_*xk*_ = 0 for all the other clusters. When there are two outcomes, let *q*_*y*1_ = 0.4 and *q*_*y*2_ = 0.1. The causal effect parameters are set as *θ*_*k*1_ = (0.1, 0.3, 0.5, 0.4) for the first outcome, and *θ*_*k*2_ = (0.2, *−*0.3, 0.6, 0) for the second outcome, with *k* = 1, …, 4. Causal effects of the 10 noise variants are generated from *N* (0, 1). When there are three outcomes, additionally set *q*_*y*3_ = 0.2 and *θ*_*k*3_ = (*−*0.2, 0.3, 0, 0.3).

When there is *K* = 1 substantive cluster (with 90 variants) and one noise cluster (with 10 variants), set 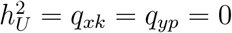 and 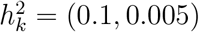. The causal effects of *X* on all the outcomes are set to zero. In all simulation designs, *δ*_*kj*_ and *η*_*j*_ are generated from the uniform distribution *U* [0.1, 0.3], and are randomly assigned to be positive or negative, then re-scaled as 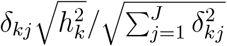 and 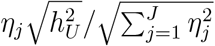 to make sure that variations in *U* and *X*_*k*_ explained by the variants equal to 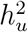 and 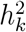 respectively.

The Rand index [66] is a quantity which measures the similarity between two clustering outcomes with values between 0 and 1. It is given by 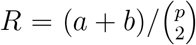. Here, *a* denotes the number of pairs of objects that are classified as belonging to the same cluster in both clustering outcomes and *b* is the number of pairs of objects that are classified in different clusters by both clustering outcomes. *R* values close to 1 indicate good agreement and values close to 0 indicate poor agreement between two clustering outcomes. Here the Rand index is calculated with variants assigned to the substantive clusters by the methods. The mean absolute error (MAE) and mean squared error (MSE) are calculated as follows:

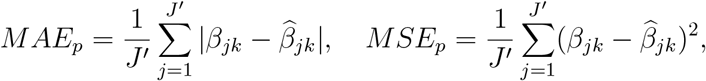

where *J*^*′*^ is the number of variants that are not assigned to the junk cluster by the methods, and do not belong to the junk cluster by the ground truth. *β*_*jk*_ is the true causal effect associated with the cluster which *G*_*j*_ truly belongs to, and 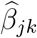 is the causal estimate associated with the cluster which *G*_*j*_ is assigned to by the methods. The subscript *p* denotes the *p*-th outcome, and the overall MAE and MSE are calculated as means over all the outcomes.

MR-AHC is performed with the outlier-robust variation as described previously. To avoid spurious clusters, we only report the detected clusters containing more than 4 variants. Small clusters with less than 4 variants are subsumed into the junk cluster. The inputs for NAvMix are standardized before being supplied to the algorithm, as recommended in Grant et al. [25]. The cluster-specific causal estimates are obtained using the IVW approach. One exception is that if there is overdispersion within a detected cluster, as indicated by a non-zero *I*^2^, then the cluster-specific estimate and its standard error are calculated using MR-RAPS [13] to account for the within-cluster overdispersion.

### Clustering analysis on the BFP associated genetic variants based on the causal estimates of T2D and OA

We use SNP-BFP summary data from a GWAS study based on UK Biobank individuals from Martin et al. [24], including 696 SNPs at genome-wide significance (*p <* 5 *×* 10^*−*8^). The T2D GWAS statistics are from Mahajan et al. [67], which combine 31 published GWAS studies but exclude the UK Biobank individuals. The SNP-OA summary statistics are from a FinnGen GWAS (code: M13_ARTHROSIS_INCLAVO) [68]. Only SNPs present in all three datasets are used for analyses (487 in total). SNPs are orientated across all three datasets in the direction of increasing the exposure. The T2D and OA samples are non-overlapping, therefore for each SNP, the covariance between the SNP-T2D estimate and SNP-OA estimate is treated as zero.

In implementing the MR-AHC method, we use the effective sample size [30] of the T2D GWAS sample (*n* = 193, 440) to calculate the threshold p-value 0.1*/* log(*n*) in the binary outcome setting for Cochran’s Q test and the post-selection Wald test in detecting the null clusters. Clustering results of MR-AHC are obtained using an iterated outlier removal procedure: this performs the outlier removal and re-fitting indefinitely until the individual p-values of the Q statistics for all SNPs are above 5%. The cluster-specific causal estimates and standard errors are calculated with the IVW approach. For clusters with overdispersion indicated by a non-zero *I*^2^, the estimates are obtained using MR-RAPS to account for the within-cluster overdispersion. We set the initial proportion of noise SNPs as 5% for both mclust and NAvMix.

### Post-clustering analysis

We map SNPs in each cluster to genes using the SNP2GENE function in FUMA based on positional mapping (with deleterious coding SNPs) [69], eQTL mapping, and chromatin interaction mapping. This three-way mapping strategy is used in the applied examples in the original paper introducing FUMA [32]. The uploaded SNPs are also set to be the pre-defined lead SNPs. All default settings are applied, with the exception that we set the reference panel population as “UKB release2b 10k European”. For eQTL mapping, following the practice in Grant et al. [25], we select tissue types from the following data sources: eQTL catalogue, PsychENCODE, van der Wijst et al. scRNA eQTLs, DICE, eQTLGen, Blood eQTLs, MuTHER, xQTLServer, ComminMind Consortium, BRAINEAC and GTEx v8 [70–79]. For chromatin interaction mapping, we select all available Hi-C datasets. The gene-set enrichment analysis is conducted using the GENE2FUNC function in FUMA. For the mapped genes corresponding to each cluster, we perform the hypergeometric test to check if the mapped genes are over-represented in a pre-defined gene set. Multi-testing correction with the Benjamini-Hochberg procedure is applied, with the adjusted p-value *≤* 0.05 as threshold [32]. The pre-defined gene sets for canonical pathways and gene ontology processes are obtained from MsigDB and WikiPathways [80, 81]. Gene sets for phenotypes are from GWAS catalog [82].

To test how the variant clusters are associated with oxidative stress, we create a list of 11 OS biomarkers from various categories, including: glutathione transferase (GST), catalase(CAT), superoxide dismutase(SOD), glutathione peroxidase (GPX), C-reactive protein (CRP), Interleukin 6 (IL-6), Tumor necrosis factor alpha (TNF-*α*), Interleukin 1 beta (IL-1*β*), Interleukin 12 (IL-12), Interleukin 8 (IL-8) and Growth/differentiation factor-15 (GDF-15). The GWAS summary statistics for the four antioxidants (GST, CAT, SOD, GPX) and CRP are obtained from the GWAS of Sun et al. [83]; for GDF-15, the GWAS of Gudjonsson et al. [84]; for the rest five cytokines, the GWAS of Ahola-Olli et al. and Kalaoja et al. [86]. Summary statistics for bipolar disorder and major depression disorder are taken from two GWAS studies conducted by the Psychiatric Genomics Consortium [87, 88]. For MR analyses associated with Cluster 4, the WHR (adjust for BMI) statistics are obtained from a GWAS conducted by the GIANT Consortium [89]; for the HDL-C and total cholesterol data, the GWAS from the Global Lipids Genetics Consortium [90]; for CAD, the GWAS from the CARDIoGRAMplusC4D Consortium [91].

## Data Availability

All data used in the applied examples in the present study are available upon reasonable request to the authors. The R code that generates the simulation datasets are available online at https://github.com/xiaoran-liang/MRAHC

## Funding

This work was supported by the Strategic Priority Fund “Tackling multimorbidity at scale” programme (grant number MC/MR/WO14548/1) delivered by the Medical Research Council and the National Institute for Health and Care Research in partnership with the Economic and Social Research Council and in collaboration with the Engineering and Physical Sciences Research Council. Nicolas Apfel is supported by the ESRC grant EST013567/1.

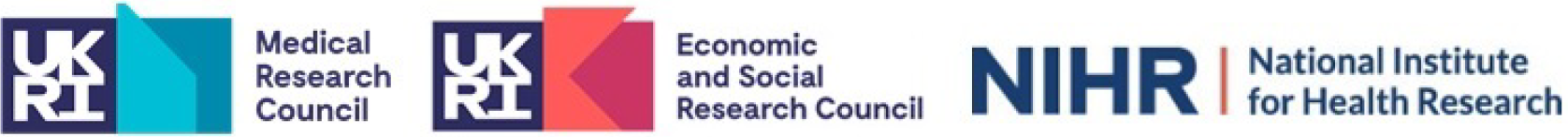

