## Appendix for "Using clustering of genetic variants in Mendelian randomization to interrogate the causal pathways underlying multimorbidity"

### A The weighted squared Euclidean distance

Without loss of generality, consider two variant clusters $\mathcal{S}_k$ and $\mathcal{S}_l$. Suppose the true underlying causal effect from the exposure to the outcome $Y_p$ identified by $\mathcal{S}_k$ is $\beta_{\mathcal{S}_k,p}$. We refer to $\beta_{\mathcal{S}_k,p}$ as the causal estimand of $\mathcal{S}_k$. Similarly, let $\beta_{\mathcal{S}_l,p}$ be the causal estimand of $\mathcal{S}_l$. In all the relevant notations, $p = 1, ..., P$. To test if $\mathcal{S}_k$ and $\mathcal{S}_l$ identify the same causal effects across all outcomes, i.e. to test the null hypothesis

$$H_0 : \boldsymbol{\beta}_{\mathcal{S}_k} = \boldsymbol{\beta}_{\mathcal{S}_l}$$

where $\boldsymbol{\beta}_{\mathcal{S}_k}$, $\boldsymbol{\beta}_{\mathcal{S}_l}$ are P-dimensional vectors of $\beta_{\mathcal{S}_k,p}$ and $\beta_{\mathcal{S}_l,p}$ respectively, we can construct the Wald statistic using the IVW estimates of $\boldsymbol{\beta}_{\mathcal{S}_k}$ and $\boldsymbol{\beta}_{\mathcal{S}_l}$, denoted by $\widehat{\boldsymbol{\beta}}^{IVW}_{\boldsymbol{\mathcal{S}_k}}$ and $\widehat{\boldsymbol{\beta}}^{IVW}_{\boldsymbol{\mathcal{S}_l}}$, as defined in Equation (10), with the $p$-th entry

$$\widehat{\beta}^{IVW}_{\mathcal{S}_k,p} = \frac{\sum_{j\in S_k} \widehat{\beta}_{jp} w_{jp}}{\sum_{j\in S_k} w_{jp}}.$$

Then the Wald testing statistic for the above null hypothesis is defined as

$$\mathcal{W}_{k,l} = \left(\widehat{\boldsymbol{\beta}}^{IVW}_{\boldsymbol{\mathcal{S}_k}} - \widehat{\boldsymbol{\beta}}^{IVW}_{\boldsymbol{\mathcal{S}_l}}\right)' \widehat{\boldsymbol{Var}}^{-1}_{k,l} \left(\widehat{\boldsymbol{\beta}}^{IVW}_{\boldsymbol{\mathcal{S}_k}} - \widehat{\boldsymbol{\beta}}^{IVW}_{\boldsymbol{\mathcal{S}_l}}\right), \tag{S1}$$

where $\widehat{\boldsymbol{Var}}_{k,l}$ is the covariance matrix of $\left(\widehat{\boldsymbol{\beta}}^{IVW}_{\boldsymbol{\mathcal{S}_k}} - \widehat{\boldsymbol{\beta}}^{IVW}_{\boldsymbol{\mathcal{S}_l}}\right)$. The diagonal entries of $\widehat{\boldsymbol{Var}}_{k,l}$ are the variances of $\left(\widehat{\beta}^{IVW}_{\mathcal{S}_k,p} - \widehat{\beta}^{IVW}_{\mathcal{S}_l,p}\right)$. We assume that all the variants are independent of each other, and all the ratio estimates across different variants are uncorrelated, then we have

$$Var\left(\widehat{\beta}^{IVW}_{\mathcal{S}_k,p} - \widehat{\beta}^{IVW}_{\mathcal{S}_l,p}\right) = Var(\widehat{\beta}^{IVW}_{\mathcal{S}_k,p}) + Var(\widehat{\beta}^{IVW}_{\mathcal{S}_l,p}).$$

The variance of $\widehat{\beta}^{IVW}_{\mathcal{S}_k,p}$ is given by

$$Var(\widehat{\beta}^{IVW}_{\mathcal{S}_k,p}) = \frac{1}{W^2_{kp}} \sum_{j\in\mathcal{S}_k} w^2_{jp} Var(\widehat{\beta}_{jp}) = \frac{1}{W^2_{kp}} \sum_{j\in\mathcal{S}_k} w^2_{jp} \frac{1}{w_{jp}} = \frac{1}{W_{kp}}$$

where $W_{kp} = \sum_{j\in\mathcal{S}_k} w_{jp}$ and $w_{jp} = 1/Var(\widehat{\beta}_{jp})$. Then we have

$$Var\left(\widehat{\beta}^{IVW}_{\mathcal{S}_k,p} - \widehat{\beta}^{IVW}_{\mathcal{S}_l,p}\right) = \frac{1}{W_{kp}} + \frac{1}{W_{lp}}. \tag{S2}$$

Thus far, we have the diagonal entries of $\widehat{\boldsymbol{Var}}_{k,l}$ as $\left(\frac{1}{W_{kp}} + \frac{1}{W_{lp}}\right)$ with $p = 1, ..., P$. Now we need to derive the covariance term between $\left(\widehat{\beta}^{IVW}_{\mathcal{S}_k,i} - \widehat{\beta}^{IVW}_{\mathcal{S}_l,i}\right)$ and $\left(\widehat{\beta}^{IVW}_{\mathcal{S}_k,r} - \widehat{\beta}^{IVW}_{\mathcal{S}_l,r}\right)$, with $i, r \in \{1, ..., P\}$, $i \neq r$. Start with

$$\begin{aligned}
&cov\left(\left(\widehat{\beta}^{IVW}_{\mathcal{S}_k,i} - \widehat{\beta}^{IVW}_{\mathcal{S}_l,i}\right), \left(\widehat{\beta}^{IVW}_{\mathcal{S}_k,r} - \widehat{\beta}^{IVW}_{\mathcal{S}_l,r}\right)\right) \\
&= cov\left(\widehat{\beta}^{IVW}_{\mathcal{S}_k,i}, \widehat{\beta}^{IVW}_{\mathcal{S}_k,r}\right) + cov\left(\widehat{\beta}^{IVW}_{\mathcal{S}_l,r}, \widehat{\beta}^{IVW}_{\mathcal{S}_l,r}\right) \\
&= cov\left(\frac{\sum_{j\in\mathcal{S}_k} \widehat{\beta}_{ji} w_{ji}}{\sum_{j\in\mathcal{S}_k} w_{ji}}, \frac{\sum_{j\in\mathcal{S}_k} \widehat{\beta}_{jr} w_{jr}}{\sum_{j\in\mathcal{S}_k} w_{jr}}\right) + cov\left(\frac{\sum_{j\in\mathcal{S}_l} \widehat{\beta}_{ji} w_{ji}}{\sum_{j\in\mathcal{S}_l} w_{ji}}, \frac{\sum_{j\in\mathcal{S}_l} \widehat{\beta}_{jr} w_{jr}}{\sum_{j\in\mathcal{S}_l} w_{jr}}\right)
\end{aligned} \tag{S3}$$

For the first term, we have

$$\begin{aligned}
&cov\left(\frac{\sum_{j\in\mathcal{S}_k} \widehat{\beta}_{ji} w_{ji}}{\sum_{j\in\mathcal{S}_k} w_{ji}}, \frac{\sum_{j\in\mathcal{S}_k} \widehat{\beta}_{jr} w_{jr}}{\sum_{j\in\mathcal{S}_k} w_{jr}}\right) \\
&\approx \frac{1}{W_{ki}W_{kr}} cov\left(\sum_{j\in\mathcal{S}_k} \widehat{\beta}_{ji} w_{ji}, \sum_{j\in\mathcal{S}_k} \widehat{\beta}_{jr} w_{jr}\right) \\
&\approx \frac{1}{W_{ki}W_{kr}} \sum_{j\in\mathcal{S}_k} w_{ji} w_{jr} cov(\widehat{\beta}_{ji}, \widehat{\beta}_{jr})
\end{aligned} \tag{S4}$$

Now we need to derive $cov(\widehat{\beta}_{ji}, \widehat{\beta}_{jr})$. By definition, we have

$$cov(\widehat{\beta}_{ji}, \widehat{\beta}_{jr}) = cov\left(\frac{\widehat{\Gamma}_{ji}}{\widehat{\gamma}_j}, \frac{\widehat{\Gamma}_{jr}}{\widehat{\gamma}_j}\right) \approx \frac{cov\left(\widehat{\Gamma}_{ji}, \widehat{\Gamma}_{jr}\right)}{\widehat{\gamma}_j^2}$$

Consider a case where we obtain $\left(\widehat{\Gamma}_{ji}, \widehat{\Gamma}_{jr}, \widehat{\gamma}_j\right)$ from three samples with *i.i.d.* individuals, we have

$$Y_i = G_j \Gamma_{ji} + \epsilon_{ji}$$

$$Y_r = G_j \Gamma_{jr} + \epsilon_{jr}$$

where $G_j$ is independent with $\epsilon_{ji}$ and $\epsilon_{jr}$. Assume $Y_i$, $Y_r$ and $G_j$ all have mean 0, and $G_j$ has variance equal to 1. Let $N_i$, $N_r$ and $N_o$ denote the sample size for $Y_i$, $Y_r$ and their overlap respectively. Following the derivation in Wang et al. [31] Appendix S2, we have:

$$\widehat{\Gamma}_{ji} = \frac{\widehat{cov_{N_i}(Y_i, G_j)}}{\widehat{cov_{N_i}(G_j, G_j)}} = \widehat{cov_{N_i}(Y_i, G_j)} = \frac{1}{N_i} \sum_{m=1}^{N_i} Y_{im} G_{jm}$$

$$\widehat{\Gamma}_{jr} = \frac{\widehat{cov_{N_r}(Y_r, G_j)}}{\widehat{cov_{N_r}(G_j, G_j)}} = \widehat{cov_{N_r}(Y_r, G_j)} = \frac{1}{N_r} \sum_{m=1}^{N_r} Y_{rm} G_{jm}.$$

Then

$$\begin{aligned}
cov\left(\widehat{\Gamma}_{ji}, \widehat{\Gamma}_{jr}\right) &= cov\left(\frac{1}{N_i} \sum_{m=1}^{N_i} Y_{im} G_{jm}, \frac{1}{N_r} \sum_{m=1}^{N_r} Y_{rm} G_{jm}\right) \\
&= \frac{1}{N_i N_r} cov\left(\sum_{m=1}^{N_i} Y_{im} G_{jm}, \sum_{m=1}^{N_r} Y_{rm} G_{jm}\right) \\
&= \frac{1}{N_i N_r} cov\left(\sum_{m=1}^{N_o} Y_{im} G_{jm}, \sum_{m=1}^{N_o} Y_{rm} G_{jm}\right) \\
&= \frac{N_o}{N_i N_r} cov\left(Y_i G_j, Y_r G_j\right) = \frac{N_o}{N_i N_r} cov\left(G_r^2 \Gamma_{ji} + G_j \epsilon_{ji}, G_j^2 \Gamma_{jr} + G_j \epsilon_{jr}\right) \\
&= \frac{N_o}{N_i N_r} \left(\Gamma_{ji} \Gamma_{jr} Var(G_j^2) + \Gamma_{ji} cov(G_j^2, G_j \epsilon_{ji}) + \Gamma_{jr} cov(G_j^2, G_j \epsilon_{jr}) + cov(G_j \epsilon_{ji}, G_j \epsilon_{jr})\right)
\end{aligned}$$

Following the argument in Wang et al. [31], for most the variants, their individual genetic effects $\Gamma_{ji}$ and $\Gamma_{jr}$ are very small, so approximately

$$\begin{aligned}
cov\left(\widehat{\Gamma}_{ji}, \widehat{\Gamma}_{jr}\right) &\approx \frac{N_o}{N_i N_r} cov\left(G_j \epsilon_{ji}, G_j \epsilon_{jr}\right) \\
&= \frac{N_o}{N_i N_r} \left(E(G_j^2 \epsilon_{ji} \epsilon_{ji}) - E(G_j \epsilon_{jr}) E(G_j \epsilon_{jr})\right)
\end{aligned}$$

As

$$E(G_j\epsilon_{ji}) = E_G\left(E(G_j\epsilon_{ji}|G_j)\right) = E_G\left(G_jE(\epsilon_{ji}|G_j)\right) = 0,$$

then

$$cov\left(\widehat{\Gamma}_{ji}, \widehat{\Gamma}_{jr}\right) = \frac{N_o}{N_iN_r}E(G_j^2\epsilon_{ji}\epsilon_{jr})$$

Because $G_j$ and $\epsilon_{ji}, \epsilon_{jr}$ are independent, then

$$\begin{aligned} cov\left(\widehat{\Gamma}_{ji}, \widehat{\Gamma}_{jr}\right) &= \frac{N_o}{N_iN_r}E(G_j^2\epsilon_{ji}\epsilon_{jr}) = \frac{N_o}{N_iN_r}E(G_j^2)E(\epsilon_{ji}\epsilon_{jr}) \\ &= \frac{N_o}{N_iN_r}Var(G_j^2)cov(\epsilon_{ji}\epsilon_{jr}) = \frac{N_o}{N_iN_r}cov(\epsilon_{ji}\epsilon_{jr}) \\ &= \frac{N_o}{N_iN_r}cov(Y_i - G_j\Gamma_{ji}, Y_r - G_j\Gamma_{jr}) \\ &\approx \frac{N_o}{N_iN_r}cov(Y_i, Y_r) \end{aligned}$$

By definition, the correlation between $\widehat{\Gamma}_{ji}$ and $\widehat{\Gamma}_{jr}$, denoted by $\rho_{jir}$, is

$$\rho_{jir} = \frac{cov\left(\widehat{\Gamma}_{ji}, \widehat{\Gamma}_{jr}\right)}{se\left(\widehat{\Gamma}_{ji}\right)se\left(\widehat{\Gamma}_{jr}\right)}.$$

By

$$\begin{aligned} Var(\widehat{\Gamma}_{ji}) &= Var(\frac{1}{N_i}\sum_{m=1}^{N_i}Y_{im}G_{jm}) = \frac{1}{N_i}Var(Y_iG_j) \\ &= \frac{1}{N_i}Var(G_j^2\widehat{\Gamma}_{ji} + G_j\epsilon_{ji}) \\ &\approx \frac{1}{N_i}Var(G_j\epsilon_{ji}) = \frac{1}{N_i}Var(G_j)Var(\epsilon_{ji}) \\ &= \frac{1}{N_i}Var(\epsilon_{ji}) \approx \frac{1}{N_i}Var(Y_i), \end{aligned}$$

we have

$$\rho_{jir} = \frac{cov\left(\widehat{\Gamma}_{ji}, \widehat{\Gamma}_{jr}\right)}{se\left(\widehat{\Gamma}_{ji}\right) se\left(\widehat{\Gamma}_{jr}\right)} = \frac{N_o}{\sqrt{N_i N_r}} \frac{cov(Y_i, Y_r)}{se\left(Y_i\right) se\left(Y_r\right)} = \frac{N_o}{\sqrt{N_i N_r}} corr(Y_i, Y_r),$$

where $corr(Y_i Y_r)$ is the phenotypic correlation between the two outcome traits $Y_i$ and $Y_r$. Since it is the same across $j = 1, ..., J$, we omit the subscript $j$. Given $\rho_{ir}$, we have

$$cov\left(\widehat{\Gamma}_{ji}, \widehat{\Gamma}_{jr}\right) = \rho_{ir} se\left(\widehat{\Gamma}_{ji}\right) se\left(\widehat{\Gamma}_{jr}\right).$$

Then

$$cov(\widehat{\beta}_{ji}, \widehat{\beta}_{jr}) = cov\left(\frac{\widehat{\Gamma}_{ji}}{\widehat{\gamma}_j}, \frac{\widehat{\Gamma}_{jr}}{\widehat{\gamma}_j}\right) \approx \frac{cov\left(\widehat{\Gamma}_{ji}, \widehat{\Gamma}_{jr}\right)}{\widehat{\gamma}_j^2} = \frac{\rho_{ir} se\left(\widehat{\Gamma}_{ji}\right) se\left(\widehat{\Gamma}_{jr}\right)}{\widehat{\gamma}_j^2}.$$

Plug into Equation (S4), we have

$$\begin{aligned}
& cov\left(\frac{\sum_{j \in \mathcal{S}_k} \widehat{\beta}_{ji} w_{ji}}{\sum_{j \in \mathcal{S}_k} w_{ji}}, \frac{\sum_{j \in \mathcal{S}_k} \widehat{\beta}_{jr} w_{jr}}{\sum_{j \in \mathcal{S}_k} w_{jr}}\right) \\
&= \frac{1}{W_{ki} W_{kr}} \sum_{j \in \mathcal{S}_k} w_{ji} w_{jr} cov(\widehat{\beta}_{ji}, \widehat{\beta}_{jr}) \\
&= \frac{1}{W_{ki} W_{kr}} \sum_{j \in \mathcal{S}_k} w_{ji} w_{jr} \rho_{ir} se\left(\widehat{\Gamma}_{ji}\right) se\left(\widehat{\Gamma}_{jr}\right) / \widehat{\gamma}_j^2 \\
&= \frac{\rho_{ir}}{W_{ki} W_{kr}} \sum_{j \in \mathcal{S}_k} \frac{\widehat{\gamma}_j^2}{se\left(\widehat{\Gamma}_{ji}\right) se\left(\widehat{\Gamma}_{jr}\right)}
\end{aligned}$$

as $w_{ji} = \widehat{\gamma}_j^2 / se\left(\widehat{\Gamma}_{ji}\right)^2$. Plug this into Equation (S3), we have

$$\begin{aligned}
& cov\left(\left(\widehat{\beta}^{IVW}_{\mathcal{S}_k,i} - \widehat{\beta}^{IVW}_{\mathcal{S}_l,i}\right), \left(\widehat{\beta}^{IVW}_{\mathcal{S}_k,r} - \widehat{\beta}^{IVW}_{\mathcal{S}_l,r}\right)\right) \\
&= cov\left(\frac{\sum_{j \in \mathcal{S}_k} \widehat{\beta}_{ji} w_{ji}}{\sum_{j \in \mathcal{S}_k} w_{ji}}, \frac{\sum_{j \in \mathcal{S}_k} \widehat{\beta}_{jr} w_{jr}}{\sum_{j \in \mathcal{S}_k} w_{jr}}\right) + cov\left(\frac{\sum_{j \in \mathcal{S}_l} \widehat{\beta}_{ji} w_{ji}}{\sum_{j \in \mathcal{S}_l} w_{ji}}, \frac{\sum_{j \in \mathcal{S}_l} \widehat{\beta}_{jr} w_{jr}}{\sum_{j \in \mathcal{S}_l} w_{jr}}\right) \\
&= \frac{\rho_{ir}}{W_{ki} W_{kr}} \sum_{j \in \mathcal{S}_k} \frac{\widehat{\gamma}_j^2}{se\left(\widehat{\Gamma}_{ji}\right) se\left(\widehat{\Gamma}_{jr}\right)} + \frac{\rho_{ir}}{W_{li} W_{lr}} \sum_{j \in \mathcal{S}_l} \frac{\widehat{\gamma}_j^2}{se\left(\widehat{\Gamma}_{ji}\right) se\left(\widehat{\Gamma}_{jr}\right)}.
\end{aligned} \quad \text{(S5)}$$

To this end, we obtain $\widehat{\boldsymbol{Var}}_{k,l}$ with the entry on the $i$-th row and $r$-th column defined in (S5). When $i = r$, S5 reduces to the variance term defined in S2.

We can see that the covariance matrix $\widehat{\boldsymbol{Var}}_{k,l}$ in the Wald statistic (S1) is equivalent to the weighted squared Euclidean distance $\mathcal{D}_{k,l}$ defined in (11). Therefore, merging the closest two clusters measured by the distance is equivalent to merging two clusters that have the highest similarity in their cluster-specific causal effects.

The covariance terms of $\mathcal{D}_{k,l}$ depend on both the extent of overlap between the $Y_i$ and $Y_r$ samples, and their phenotypic correlation. If the variant-outcome associations are measured with independent samples, and/or the outcome traits are uncorrelated, then all the covariance terms are zero. If the correlation is non-zero, according to Bulik-Sullivan et al. [28], $\rho_{ir}$ can be estimated from the intercept of the LD score regression:

$$E[z_{ji}z_{jr}] = \frac{\sqrt{N_i N_r}\rho_g}{M} l_j + \frac{\rho_Y N_o}{\sqrt{N_i N_r}}$$

where $z_{ji}$, $z_{jr}$ are the z-scores, $\rho_g$ is the genetic covariance, $M$ is the number of variants, and $l_j$ is the LD score, $\rho_Y$ is the phenotypic correlation between $Y_i$ and $Y_r$.

### B The threshold p-value of Cochran's Q test

We consider the case of one outcome and omit the subscript $p$ representing the $p$-th outcome in the following notations. In the downward testing procedure, define $Q_{fg}$ to be the $Q$ statistic associated with cluster $\mathcal{S}_g$ at step $f$. Also define $T_{fg}$ to be the $(1-\zeta)$ threshold of a $\chi^2$ distribution on $|\mathcal{S}_g| - 1$ degrees of freedom where $|\mathcal{S}_g|$ is the number of variants in cluster $\mathcal{S}_g$. Following a similar strategy as in Windmeijer et al. [92] and Apfel and Liang [17], we show that with $\zeta = 0.1/\log(n)$ where $n$ is the size of the sample from which we obtain the variant-outcome summary statistics $\left\{\widehat{\Gamma}_j, se\left(\widehat{\Gamma}_j\right)\right\}_{j=1}^{J}$, both the Type I and Type II error of the test associated with $Q_{fg}$ goes to 0 as $n \to \infty$.

First, we establish the asymptotic property of $Q_{fg}$ as $n \to \infty$:

1. $Q_{fg} \xrightarrow{d} \chi^2_{|\mathcal{S}_g|-1}$ if all variants in $\mathcal{S}_g$ identify the same causal effect [8].

2. $Q_{fg} = O_p(n)$ if variants in $\mathcal{S}_g$ identify different causal effects.

The proof for Point 2 is as follows (all the summations are for $j \in \mathcal{S}_g$):
Based on the definition of $\widehat{\beta}_j$ and $w_j$, by the continuous mapping theorem, we haves

$$plim(\widehat{\beta}_j) = plim(\widehat{\Gamma}_j/\widehat{\gamma}_j) = \Gamma_j/\gamma_j = \beta_j,$$

$$plim(w_j) = plim\left(1/\widehat{Var(\widehat{\beta}_j)}\right) = plim\left(\frac{\widehat{\gamma}_j^2}{se\left(\widehat{\Gamma}_j\right)^2}\right) = \frac{n\gamma_j^2}{\sigma_{Yj}^2}.$$

Then

$$\begin{aligned}
plim(Q_{fg}) &= plim\left(\sum w_j\left(\widehat{\beta}_j - \frac{\sum w_j\widehat{\beta}_j}{\sum w_j}\right)^2\right) = \sum plim\left(w_j\left(\widehat{\beta}_j - \frac{\sum w_j\widehat{\beta}_j}{\sum w_j}\right)^2\right)\\
&= \sum\left(\frac{n\gamma_j^2}{\sigma_{Yj}^2} \times plim\left(\widehat{\beta}_j - \frac{\sum w_j\widehat{\beta}_j}{\sum w_j}\right)^2\right) = \sum\left(\frac{n\gamma_j^2}{\sigma_{Yj}^2} \times plim\left(\frac{\widehat{\beta}_j\sum w_j - \sum w_j\widehat{\beta}_j}{\sum w_j}\right)^2\right)\\
&= \sum\left(\frac{n\gamma_j^2}{\sigma_{Yj}^2} \times plim\left(\frac{\widehat{\beta}_j\sum\frac{n\gamma_j^2}{\sigma_{Yj}^2} - \sum\frac{n\gamma_j^2}{\sigma_{Yj}^2}\widehat{\beta}_j}{\sum\frac{n\gamma_j^2}{\sigma_{Yj}^2}}\right)^2\right)\\
&= \sum\left(\frac{n\gamma_j^2}{\sigma_{Yj}^2} \times plim\left(\frac{\widehat{\beta}_j\sum\frac{\gamma_j^2}{\sigma_{Yj}^2} - \sum\frac{\gamma_j^2}{\sigma_{Yj}^2}\widehat{\beta}_j}{\sum\frac{\gamma_j^2}{\sigma_{Yj}^2}}\right)^2\right)\\
&= \sum\left(\frac{n\gamma_j^2}{\sigma_{Yj}^2} \times \left(\frac{\beta_j\sum\frac{\gamma_j^2}{\sigma_{Yj}^2} - \sum\frac{\gamma_j^2}{\sigma_{Yj}^2}\beta_j}{\sum\frac{\gamma_j^2}{\sigma_{Yj}^2}}\right)^2\right).
\end{aligned}$$

For $\beta_j\sum\frac{\gamma_j^2}{\sigma_{Yj}^2} - \sum\frac{\gamma_j^2}{\sigma_{Yj}^2}\beta_j = \sum_{i\neq j}(\beta_j - \beta_i)\frac{\gamma_i^2}{\sigma_{Yi}^2}$, as variants in $\mathcal{S}_g$ identify different effects, there is at least one $\beta_j - \beta_i \neq 0$. Therefore, we have $\beta_j\sum\frac{\gamma_j^2}{\sigma_{Yj}^2} - \sum\frac{\gamma_j^2}{\sigma_{Yj}^2}\beta_j \neq 0$ considering the general case where the non-zero terms do not cancel out. It follows that

$$plim(Q_{fg}) = \sum \left( \frac{n\gamma_j^2}{\sigma_{Yj}^2} \times \left( \frac{\beta_j \sum \frac{\gamma_j^2}{\sigma_{Yj}^2} - \sum \frac{\gamma_j^2}{\sigma_{Yj}^2}\beta_j}{\sum \frac{\gamma_j^2}{\sigma_{Yj}^2}} \right)^2 \right) = \sum O_p(n) = O_p(n).$$

If $T_{fg}$ satisfies $T_{fg} \to \infty$, $T_{fg} = o(n)$, it follows that

$$\lim_{n\to\infty} P\left(T_{fg} < Q_{fg}\right) = 1,$$

hence the Q test is rejected if variants in $\mathcal{S}_g$ identify different effects, and

$$\lim_{n\to\infty} P\left(Q_{fg} < T_{fg}\right) = 1,$$

if variants in $\mathcal{S}_g$ identify the same effect. In this way, both the Type I and Type II error of Cochran's Q test goes to 0 as $n \to \infty$. Andrews [62] show that $T_{fg}$ that satisfies $T_{fg} \to \infty$ and $T_{fg} = o(n)$ can be translated into the threshold p-value that satisfies $\zeta \to 0$ and $log(\zeta) = o(n)$. We then set $\zeta = 0.1/log(n)$ following the suggestion of Belloni et al. [29]. When there are multiple outcomes, the Q statistic defined in (12) is a direct extension of the following single-outcome Q:

$$Q_{fg} = \sum_{j\in\mathcal{S}_{fg}} w_j(\hat{\beta}_j - \hat{\beta}^{IVW}_{\mathcal{S}_{fg}})^2 = \left(\widehat{\boldsymbol{\beta}}_{\mathcal{S}_{fg}} - \boldsymbol{\iota}_k\hat{\beta}^{IVW}_{\mathcal{S}_{fg}}\right)' Var(\widehat{\boldsymbol{\beta}}_{fg})^{-1}\left(\widehat{\boldsymbol{\beta}}_{\mathcal{S}_{fg}} - \boldsymbol{\iota}_k\hat{\beta}^{IVW}_{\mathcal{S}_{fg}}\right),$$

where $\widehat{\boldsymbol{\beta}}_{\mathcal{S}_{fg}}$ is replaced by the $P \times |\mathcal{S}_{fg}|$ length vector $\boldsymbol{\mathcal{B}}_{\mathcal{S}_{fg}}$ combing ratio estimates across all $P$ outcomes, and $\hat{\beta}^{IVW}_{\mathcal{S}_{fg}}$ is adjusted as the IVW estimates for each outcome. In this case, the degrees of freedom would be $P \times (|\mathcal{S}_{fg}| - 1)$.

### C Direct causality between outcomes

We illustrate how the direct causality between outcomes impacts the clustering of the variants associated with the exposure. Consider the extended model, with two outcomes and three exposure sub-components, as depicted in the following DAG, in which we add direct causal effects between $Y_1$ and $Y_2$: $\lambda_1$ denotes the direct effect of $Y_1$ on $Y_2$, and $\lambda_2$ denotes the direct effect of $Y_2$ on $Y_1$. $G_1$ represents the pleiotropic variant. Maintain the assumption that all the variants are independent with each other. For ease of illustration, we assume that there is one variant in each cluster.

We then have

$$
\begin{aligned}
U &= \eta G_1 + \epsilon_U, \\
X &= X_1 + X_2 + X_3 \\
&= q_x \eta G_1 + \delta_2 G_2 + \delta_3 G_3 + \epsilon_x, \\
Y_1 &= q_{y_1} U + \theta_{21} X_2 + \theta_{31} X_3 + \lambda_2 Y_2 + \epsilon_{y_1}, \\
Y_2 &= q_{y_2} U + \theta_{22} X_2 + \theta_{32} X_3 + \lambda_1 Y_1 + \epsilon_{y_2}.
\end{aligned}
$$

where $G$ is uncorrelated with all the error terms. To express $Y_1$ and $Y_2$ only in $G$, we need to solve the following equation system:

$$
\begin{aligned}
Y_1 &= q_{y_1} U + \theta_{21} X_2 + \theta_{31} X_3 + \lambda_2 Y_2 + \epsilon_{y_1}, \\
Y_2 &= q_{y_2} U + \theta_{22} X_2 + \theta_{32} X_3 + \lambda_1 Y_1 + \epsilon_{y_2}.
\end{aligned}
$$

Plug $U$ and $X$ expressed in $G$ into the above equations, and solve the system, we then obtain

$$
\begin{aligned}
Y_1 &= \frac{q_{y_1}\eta + \lambda_2 q_{y_2}\eta}{1-\lambda_1\lambda_2} G_1 + \frac{(\lambda_2\theta_{22} + \theta_{21})\delta_2}{1-\lambda_1\lambda_2} G_2 + \frac{(\lambda_2\theta_{32} + \theta_{31})\delta_3}{1-\lambda_1\lambda_2} G_3 + \xi_1, \\
Y_2 &= \frac{q_{y_2}\eta + \lambda_1 q_{y_1}\eta}{1-\lambda_1\lambda_2} G_1 + \frac{(\lambda_1\theta_{21} + \theta_{22})\delta_2}{1-\lambda_1\lambda_2} G_2 + \frac{(\lambda_1\theta_{31} + \theta_{32})\delta_3}{1-\lambda_1\lambda_2} G_3 + \xi_2,
\end{aligned}
$$

where $G$ is uncorrelated with the implicitly defined error terms $\xi_1$, $\xi_2$. As $X$ can be expressed in $G$ as follows:

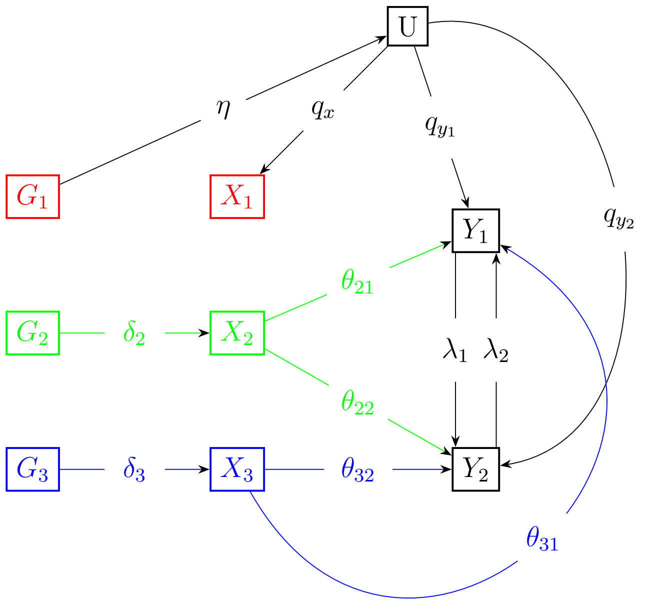


Figure S1: DAG of the extended model incorporating direct causality between outcomes.

$$X = q_x \eta G_1 + \delta_2 G_2 + \delta_3 G_3 + \epsilon_x,$$

then the causal effects identified by each variant are:

$$\begin{aligned} G_1 &: \left( \frac{q_{y_1} + \lambda_2 q_{y_2}}{(1-\lambda_1\lambda_2) q_x}, \ \frac{q_{y_2} + \lambda_1 q_{y_1}}{(1-\lambda_1\lambda_2) q_x} \right) \\ G_2 &: \left( \frac{\lambda_2 \theta_{22} + \theta_{21}}{1-\lambda_1\lambda_2}, \ \frac{\lambda_1 \theta_{21} + \theta_{22}}{1-\lambda_1\lambda_2} \right) \\ G_3 &: \left( \frac{\lambda_2 \theta_{32} + \theta_{31}}{1-\lambda_1\lambda_2}, \ \frac{\lambda_1 \theta_{31} + \theta_{32}}{1-\lambda_1\lambda_2} \right). \end{aligned} \tag{S6}$$

Since $G$ is uncorrelated with all the error terms in the aforementioned equations, and all variants are independent with each other, the variant-exposure and variant-outcome associations can be estimated consistently by regressing $X$ and $Y$ on each $G$ respectively. It follows that the ratio estimates of the variants converges to the quantities defined in (S6) as the sample size tends to infinity. Therefore, taking the outcome causality into account, variants impacting the exposure and outcomes through different pathways can still be grouped into the corresponding clusters, as only the variants belonging to the same pathway identify the same effects. However, in this case, the effects identified by each cluster would be the total effects incorporating the outcome causality, rather than the direct causal effects from the exposure to outcomes. Further work is required to address this issue.

### D Further simulation results

Table S1: Simulation results for designs with three outcomes. All methods are conducted treating the outcome correlations as 0. Statistics are calculated as the mean over 1000 replications.

| | MR-AHC | Mclust | Mclust noise | NavMix | NavMix ratio |
|---|---|---|---|---|---|
| **Three outcomes, k = 4, correlation = 0** | | | | | |
| # clusters | 4.135 | 4.379 | 3.577 | 4.904 | 3.589 |
| Rand index | 0.942 | 0.892 | 0.865 | 0.773 | 0.861 |
| # junk | 14.086 | 0.000 | 9.996 | 4.122 | 5.064 |
| # correct junk | 9.404 | 0.000 | 9.169 | 3.138 | 4.351 |
| MAE | 0.084 | 0.101 | 0.102 | 0.112 | 0.100 |
| MSE | 0.029 | 0.036 | 0.036 | 0.040 | 0.038 |
| **Three outcomes, k = 4, correlation = 0.2** | | | | | |
| # clusters | 4.124 | 4.434 | 3.661 | 4.964 | 3.628 |
| Rand index | 0.944 | 0.892 | 0.869 | 0.776 | 0.862 |
| # junk | 14.310 | 0.000 | 10.098 | 4.726 | 5.060 |
| # correct junk | 9.490 | 0.000 | 9.229 | 3.409 | 4.304 |
| MAE | 0.084 | 0.101 | 0.101 | 0.111 | 0.099 |
| MSE | 0.029 | 0.036 | 0.035 | 0.039 | 0.037 |
| **Three outcomes, k = 4, correlation = 0.7** | | | | | |
| # clusters | 4.176 | 4.955 | 5.215 | 5.791 | 4.482 |
| Rand index | 0.958 | 0.944 | 0.960 | 0.798 | 0.904 |
| # junk | 14.899 | 0.000 | 9.377 | 6.807 | 4.079 |
| # correct junk | 9.516 | 0.000 | 8.586 | 4.557 | 3.329 |
| MAE | 0.083 | 0.090 | 0.084 | 0.104 | 0.091 |
| MSE | 0.028 | 0.032 | 0.029 | 0.037 | 0.033 |
| **Three outcomes, k = 1, correlation = 0** | | | | | |
| # clusters | 1.012 | 2.170 | 1.315 | 1.004 | 1.003 |
| Rand index | 0.996 | 0.901 | 0.920 | 0.818 | 0.818 |
| # junk | 13.884 | 0.000 | 11.108 | 0.130 | 0.127 |
| # correct junk | 9.829 | 0.000 | 9.519 | 0.013 | 0.019 |
| MAE | 0.011 | 0.020 | 0.015 | 0.016 | 0.017 |
| MSE | 0.000 | 0.003 | 0.001 | 0.000 | 0.000 |
| Freq.null | 0.975 | 0.815 | 0.758 | 0.998 | 0.993 |
| **Three outcomes, k = 1, correlation = 0.2** | | | | | |
| # clusters | 1.006 | 2.226 | 1.367 | 1.015 | 1.009 |
| Rand index | 0.997 | 0.888 | 0.906 | 0.814 | 0.816 |
| # junk | 13.911 | 0.000 | 11.083 | 0.594 | 0.627 |
| # correct junk | 9.896 | 0.000 | 9.500 | 0.076 | 0.072 |
| MAE | 0.011 | 0.021 | 0.016 | 0.016 | 0.017 |
| MSE | 0.000 | 0.003 | 0.001 | 0.000 | 0.001 |
| Freq.null | 0.974 | 0.784 | 0.735 | 0.997 | 0.983 |
| **Three outcomes, k = 1, correlation = 0.7** | | | | | |
| # clusters | 1.008 | 2.180 | 1.297 | 1.932 | 1.935 |
| Rand index | 0.995 | 0.916 | 0.941 | 0.535 | 0.537 |
| # junk | 13.514 | 0.000 | 10.935 | 48.183 | 48.256 |
| # correct junk | 9.940 | 0.000 | 9.687 | 8.038 | 8.096 |
| MAE | 0.012 | 0.019 | 0.014 | 0.030 | 0.132 |
| MSE | 0.000 | 0.002 | 0.001 | 0.002 | 0.019 |
| Freq.null | 0.952 | 0.801 | 0.764 | 0.869 | 0.002 |

Table S2: Simulation results for designs with two outcomes. All methods are conducted with the true correlation parameters where feasible. Statistics are calculated as the mean over 1000 replications.

| | MR-AHC | Mclust | Mclust noise | NavMix | NavMix ratio |
|---|---|---|---|---|---|
| **Two outcomes, k = 4, correlation = 0** | | | | | |
| # clusters | 4.196 | 3.715 | 2.963 | 3.018 | 2.307 |
| Rand index | 0.917 | 0.757 | 0.737 | 0.615 | 0.710 |
| # junk variants | 10.966 | 0.000 | 7.726 | 6.517 | 4.080 |
| # correct junk | 6.975 | 0.000 | 7.135 | 1.446 | 3.951 |
| MAE | 0.088 | 0.120 | 0.123 | 0.162 | 0.106 |
| MSE | 0.030 | 0.041 | 0.041 | 0.052 | 0.035 |
| **Two outcomes, k = 4, correlation = 0.2** | | | | | |
| # clusters | 4.163 | 3.665 | 2.933 | 3.074 | 2.161 |
| Rand index | 0.919 | 0.744 | 0.725 | 0.620 | 0.687 |
| # junk variants | 11.268 | 0.000 | 7.833 | 1.281 | 4.083 |
| # correct junk | 7.257 | 0.000 | 7.233 | 0.315 | 4.007 |
| MAE | 0.088 | 0.118 | 0.122 | 0.156 | 0.101 |
| MSE | 0.030 | 0.039 | 0.040 | 0.049 | 0.032 |
| **Two outcomes, k = 4, correlation = 0.7** | | | | | |
| # clusters | 4.153 | 3.652 | 3.754 | 3.784 | 2.519 |
| Rand index | 0.950 | 0.784 | 0.835 | 0.684 | 0.751 |
| # junk variants | 12.992 | 0.000 | 8.044 | 4.907 | 4.994 |
| # correct junk | 8.271 | 0.000 | 7.433 | 0.909 | 4.648 |
| MAE | 0.083 | 0.091 | 0.088 | 0.113 | 0.096 |
| MSE | 0.028 | 0.027 | 0.027 | 0.031 | 0.026 |
| **Two outcomes, k = 1, correlation = 0** | | | | | |
| # clusters | 1.026 | 2.211 | 1.312 | 1.000 | 1.001 |
| Rand index | 0.947 | 0.887 | 0.895 | 0.818 | 0.818 |
| # junk variants | 10.194 | 0.000 | 8.211 | 0.000 | 0.000 |
| # correct junk | 7.703 | 0.000 | 7.073 | 0.000 | 0.000 |
| MAE | 0.012 | 0.020 | 0.015 | 0.017 | 0.017 |
| MSE | 0.000 | 0.004 | 0.001 | 0.000 | 0.000 |
| Freq.null | 0.955 | 0.726 | 0.767 | 0.998 | 0.997 |
| **Two outcomes, k = 1, correlation = 0.2** | | | | | |
| # clusters | 1.039 | 2.226 | 1.297 | 1.001 | 1.001 |
| Rand index | 0.948 | 0.881 | 0.893 | 0.818 | 0.818 |
| # junk variants | 10.563 | 0.000 | 8.438 | 0.015 | 0.000 |
| # correct junk | 7.919 | 0.000 | 7.199 | 0.001 | 0.000 |
| MAE | 0.012 | 0.021 | 0.015 | 0.017 | 0.017 |
| MSE | 0.000 | 0.004 | 0.001 | 0.000 | 0.000 |
| Freq.null | 0.950 | 0.690 | 0.794 | 0.997 | 0.996 |
| **Two outcomes, k = 1, correlation = 0.7** | | | | | |
| # clusters | 1.008 | 2.221 | 1.296 | 1.002 | 1.001 |
| Rand index | 0.978 | 0.910 | 0.924 | 0.818 | 0.818 |
| # junk variants | 12.155 | 0.000 | 9.458 | 0.017 | 0.017 |
| # correct junk | 9.122 | 0.000 | 8.323 | 0.001 | 0.003 |
| MAE | 0.011 | 0.019 | 0.014 | 0.017 | 0.017 |
| MSE | 0.000 | 0.003 | 0.001 | 0.000 | 0.000 |
| Freq.null | 0.979 | 0.413 | 0.778 | 0.935 | 0.935 |

Table S3: Simulation results for designs with three outcomes. All methods are conducted with the true correlation parameters where feasible. Statistics are calculated as the mean over 1000 replications.

| | MR-AHC | Mclust | Mclust noise | NavMix | NavMix ratio |
|---|---|---|---|---|---|
| **Three outcomes, k = 4, correlation = 0** | | | | | |
| # clusters | 4.046 | 4.379 | 3.577 | 4.907 | 3.589 |
| Rand index | 0.948 | 0.892 | 0.865 | 0.773 | 0.861 |
| # junk | 19.616 | 0.000 | 9.996 | 4.122 | 5.064 |
| # correct junk | 9.579 | 0.000 | 9.169 | 3.140 | 4.351 |
| MAE | 0.084 | 0.101 | 0.102 | 0.112 | 0.100 |
| MSE | 0.029 | 0.036 | 0.036 | 0.040 | 0.038 |
| **Three outcomes, k = 4, correlation = 0.2** | | | | | |
| # clusters | 4.110 | 4.434 | 3.659 | 4.878 | 3.546 |
| Rand index | 0.948 | 0.892 | 0.869 | 0.774 | 0.866 |
| # junk | 14.385 | 0.000 | 10.099 | 4.533 | 5.406 |
| # correct junk | 9.575 | 0.000 | 9.228 | 3.287 | 4.561 |
| MAE | 0.084 | 0.101 | 0.101 | 0.112 | 0.098 |
| MSE | 0.029 | 0.036 | 0.035 | 0.039 | 0.037 |
| **Three outcomes, k = 4, correlation = 0.7** | | | | | |
| # clusters | 4.215 | 4.955 | 5.215 | 6.033 | 3.583 |
| Rand index | 0.981 | 0.944 | 0.960 | 0.804 | 0.898 |
| # junk | 16.019 | 0.000 | 9.377 | 5.885 | 6.328 |
| # correct junk | 9.819 | 0.000 | 8.586 | 4.255 | 5.463 |
| MAE | 0.081 | 0.090 | 0.084 | 0.100 | 0.093 |
| MSE | 0.027 | 0.032 | 0.029 | 0.030 | 0.030 |
| **Three outcomes, k = 1, correlation = 0** | | | | | |
| # clusters | 1.004 | 2.170 | 1.315 | 1.004 | 1.003 |
| Rand index | 0.997 | 0.901 | 0.920 | 0.818 | 0.818 |
| # junk | 18.908 | 0.000 | 11.108 | 0.130 | 0.127 |
| # correct junk | 9.877 | 0.000 | 9.519 | 0.013 | 0.019 |
| MAE | 0.012 | 0.020 | 0.015 | 0.016 | 0.017 |
| MSE | 0.000 | 0.003 | 0.001 | 0.000 | 0.000 |
| Freq.null | 0.978 | 0.815 | 0.758 | 0.998 | 0.993 |
| **Three outcomes, k = 1, correlation = 0.2** | | | | | |
| # clusters | 1.002 | 2.226 | 1.368 | 1.003 | 1.001 |
| Rand index | 0.998 | 0.887 | 0.906 | 0.817 | 0.818 |
| # junk | 14.044 | 0.000 | 11.080 | 0.417 | 0.011 |
| # correct junk | 9.924 | 0.000 | 9.499 | 0.037 | 0.001 |
| MAE | 0.011 | 0.021 | 0.016 | 0.016 | 0.016 |
| MSE | 0.000 | 0.003 | 0.001 | 0.000 | 0.000 |
| Freq.null | 0.979 | 0.789 | 0.739 | 0.999 | 0.997 |
| **Three outcomes, k = 1, correlation = 0.7** | | | | | |
| # clusters | 1.000 | 2.180 | 1.298 | 1.002 | 1.004 |
| Rand index | 1.000 | 0.916 | 0.941 | 0.817 | 0.818 |
| # junk | 14.205 | 0.000 | 10.936 | 0.409 | 0.098 |
| # correct junk | 9.999 | 0.000 | 9.686 | 0.024 | 0.012 |
| MAE | 0.011 | 0.019 | 0.014 | 0.016 | 0.016 |
| MSE | 0.000 | 0.002 | 0.001 | 0.000 | 0.000 |
| Freq.null | 0.958 | 0.522 | 0.771 | 0.951 | 0.947 |

| | $K=4$ | | $K=1$ | |
|---|---|---|---|---|
| | MR-AHC | MR-Clust | MR-AHC | MR-Clust |
| #clusters | 3.961 | 4.119 | 1.212 | 1.150 |
| Rand index | 0.920 | 0.918 | 0.919 | 0.898 |
| #junk | 6.018 | 2.548 | 8.484 | 4.667 |
| # correctjunk | 3.313 | 2.441 | 5.981 | 4.644 |
| MAE | 0.089 | 0.085 | 0.017 | 0.011 |
| MSE | 0.029 | 0.028 | 0.002 | 0.000 |

Table S4: Simulation results for designs with one outcome using MR-AHC and MR-Clust[15]. Data are simulated similarly as in the main text. When there are $K = 4$ substantive clusters, set $\beta = (0.1, 0.3, -0.5, 0)$ and $q_y = 0.4$. With $K = 1$, set $\beta = 0$. All other parameters are the same as in the main text. Statistics are calculated as the mean over 1000 replications.

### E Application results

| Cluster | # SNPs | Est-T2D | t-T2D | Est-OA | t-OA | $Q$ p-value | $I^2$ |
|---|---|---|---|---|---|---|---|
| 1 | 124 | 1.326 | 18.236 | 1.250 | 23.321 | 0.929 | 0.000 |
| | | (0.073) | | (0.054) | | | |
| 2 | 258 | 1.076 | 21.102 | 0.067 | 1.791 | 0.951 | 0.000 |
| | | (0.051) | | (0.037) | | | |
| 3 | 32 | −1.068 | 7.718 | 0.738 | 7.327 | 1.000 | 0.000 |
| | | (0.138) | | (0.101) | | | |
| 4 | 22 | −2.799 | 14.704 | 0.103 | 0.722 | 0.437 | 0.019 |
| | | (0.190) | | (0.143) | | | |

Table S5: The estimation results using SNPs in each cluster detected by MR-AHC for the causal relationship between BFP and T2D-OA. We report the number of SNPs in the cluster (*#SNPs*), the cluster-specific IVW estimate (*Est*), the standard error (in the parentheses under the estimates) and associated t-statistic ($t$), and statistics measuring the heterogeneity within each cluster (p-value of the Q statistic and the $I^2$ statistic). For Cluster 4 with within-cluster overdispersion indicated by $I^2 > 0$, we report the MR-RAPS estimates and standard errors correcting for overdispersion.

| Cluster | # SNPs | Est-T2D | t-T2D | Est-OA | t-OA | $Q$ p-value | $I^2$ |
|---|---|---|---|---|---|---|---|
| 1 | 411 | 1.181<br>(0.038) | 30.753 | 0.472<br>(0.028) | 16.885 | 0 | 0.416 |
| 2 | 72 | −0.823<br>(0.093) | 8.811 | 0.576<br>(0.069) | 8.367 | 0 | 0.880 |

Table S6: The estimation results using SNPs in each cluster detected by mclust for the causal relationship between BFP and T2D-OA. We report the number of SNPs in the cluster (*#SNPs*), the cluster-specific IVW estimate (*Est*), the standard error (in the parentheses under the estimates) and associated t-statistic ($t$), and statistics measuring the heterogeneity within each cluster (p-value of the Q statistic and the $I^2$ statistic).

| Cluster | # SNPs | Est-T2D | t-T2D | Est-OA | t-OA | $Q$ p-value | $I^2$ |
|---|---|---|---|---|---|---|---|
| 1 | 249 | 1.798<br>(0.047) | 37.874 | 0.606<br>(0.035) | 17.370 | 0.000 | 0.269 |
| 2 | 238 | −0.375<br>(0.053) | 7.069 | 0.363<br>(0.039) | 9.412 | 0 | 0.811 |

Table S7: The estimation results using SNPs in each cluster detected by NAvMix with zero initial noise proportion for the causal relationship between BFP and T2D-OA. We report the number of SNPs in the cluster (*#SNPs*), the cluster-specific IVW estimate (*Est*), the standard error (in the parentheses under the estimates) and associated t-statistic ($t$), and statistics measuring the heterogeneity within each cluster (p-value of the Q statistic and the $I^2$ statistic).

| | IVW | | | MR-PRESSO | | | MR-RAPS | | |
|---|---|---|---|---|---|---|---|---|---|
| | Est | SE | Z-score | Est | SE | Z-score | Est | SE | Z-score |
| GST | 0.440 | 0.216 | 2.037 | 0.440 | 0.207 | 2.126 | 0.448 | 0.221 | 2.030 |
| CAT | -0.083 | 0.216 | -0.385 | -0.083 | 0.223 | -0.373 | -0.091 | 0.227 | -0.400 |
| SOD | 0.026 | 0.216 | 0.120 | 0.026 | 0.214 | 0.121 | 0.026 | 0.221 | 0.120 |
| GPX | 0.040 | 0.216 | 0.186 | 0.040 | 0.223 | 0.181 | 0.044 | 0.226 | 0.193 |
| CRP | 0.545 | 0.167 | 3.269 | 0.545 | 0.166 | 3.275 | 0.552 | 0.178 | 3.107 |
| IL-6 | 0.345 | 0.141 | 2.449 | 0.345 | 0.148 | 2.330 | 0.351 | 0.145 | 2.415 |
| TNF-$\alpha$ | 0.200 | 0.215 | 0.929 | 0.200 | 0.215 | 0.932 | 0.204 | 0.220 | 0.927 |
| IL-1$\beta$ | 0.406 | 0.220 | 1.841 | 0.406 | 0.232 | 1.749 | 0.413 | 0.235 | 1.757 |
| IL-12 | 0.219 | 0.140 | 1.561 | 0.219 | 0.150 | 1.455 | 0.220 | 0.151 | 1.461 |
| IL-8 | 0.345 | 0.213 | 1.619 | 0.345 | 0.220 | 1.565 | 0.354 | 0.225 | 1.570 |
| GDF-15 | 0.264 | 0.150 | 1.757 | 0.264 | 0.138 | 1.911 | 0.268 | 0.154 | 1.746 |
| Bipolar | 0.232 | 0.083 | 2.812 | 0.127 | 0.123 | 1.033 | 0.226 | 0.149 | 1.518 |
| MDD | 0.114 | 0.080 | 1.425 | 0.159 | 0.102 | 1.558 | 0.135 | 0.114 | 1.183 |

Table S8: Two-sample MR estimating the effects of BFP on the 11 oxidative stress biomarkers and 2 psychological disorders using variants in Cluster 1 as instruments. Results are given by IVW, MR-PRESSO and MR-RAPS, including the point estimate ("*Est*"), the standard error ("*SE*") and the Z-score (the ratio of the estimate and the standard error).

| | IVW | | | MR-PRESSO | | | MR-RAPS | | |
|---|---|---|---|---|---|---|---|---|---|
| | Est | SE | Z-score | Est | SE | Z-score | Est | SE | Z-score |
| GST | -0.904 | 0.491 | -1.839 | -0.904 | 0.538 | -1.680 | -0.979 | 0.538 | -1.819 |
| CAT | -0.601 | 0.492 | -1.223 | -0.601 | 0.513 | -1.172 | -0.618 | 0.508 | -1.217 |
| SOD | 0.878 | 0.491 | 1.787 | 0.878 | 0.588 | 1.492 | 0.778 | 0.577 | 1.349 |
| GPX | -0.376 | 0.492 | -0.765 | -0.376 | 0.576 | -0.653 | -0.173 | 0.587 | -0.294 |
| CRP | 0.662 | 0.337 | 1.961 | 0.662 | 0.308 | 2.149 | 0.672 | 0.345 | 1.949 |
| IL-6 | -0.307 | 0.311 | -0.988 | -0.307 | 0.232 | -1.323 | -0.310 | 0.318 | -0.975 |
| TNF-$\alpha$ | -0.004 | 0.484 | -0.008 | -0.004 | 0.455 | -0.009 | -0.004 | 0.493 | -0.008 |
| IL-1$\beta$ | -0.170 | 0.495 | -0.344 | -0.170 | 0.338 | -0.504 | -0.171 | 0.507 | -0.338 |
| IL-12 | -0.923 | 0.310 | -2.981 | -0.923 | 0.243 | -3.796 | -0.934 | 0.318 | -2.937 |
| IL-8 | -0.259 | 0.477 | -0.543 | -0.259 | 0.441 | -0.588 | -0.263 | 0.487 | -0.539 |
| GDF-15 | -0.899 | 0.351 | -2.564 | -0.899 | 0.478 | -1.880 | -1.145 | 0.545 | -2.101 |
| Bipolar | 0.208 | 0.189 | 1.104 | 0.208 | 0.207 | 1.008 | 0.224 | 0.209 | 1.069 |
| MDD | 0.092 | 0.179 | 0.513 | 0.092 | 0.207 | 0.442 | 0.146 | 0.216 | 0.674 |
| WHRadj | -0.529 | 0.101 | -5.208 | -0.634 | 0.249 | -2.550 | -0.476 | 0.281 | -1.692 |
| HDL-C | 0.926 | 0.097 | 9.500 | 1.280 | 0.215 | 5.949 | 0.932 | 0.234 | 3.990 |
| TC | -0.414 | 0.101 | -4.096 | -0.192 | 0.104 | -1.853 | -0.395 | 0.139 | -2.838 |
| CAD | -0.887 | 0.416 | -2.131 | -0.887 | 0.433 | -2.049 | -0.896 | 0.425 | -2.108 |

Table S9: Two-sample MR estimating the effects of BFP on the 11 oxidative stress biomarkers, 2 psychological disorders, WHR (adjusted for BMI), HDL-C, total cholesterol (TC) and CAD using variants in Cluster 4 as instruments. Results are given by IVW, MR-PRESSO and MR-RAPS, including the point estimate ("*Est*"), the standard error ("*SE*") and the Z-score (the ratio of the estimate and the standard error).
